# Polygenic prediction of human longevity on the supposition of pervasive pleiotropy

**DOI:** 10.1101/2023.12.10.23299795

**Authors:** M.Reza Jabalameli, Jhih-Rong Lin, Quanwei Zhang, Zhen Wang, Joydeep Mitra, Nha Nguyen, Tina Gao, Mark Khusidman, Gil Atzmon, Sofiya Milman, Jan Vijg, Nir Barzilai, Zhengdong D. Zhang

**Affiliations:** Department of Genetics, Albert Einstein College of Medicine, New York, NY, USA; Department of Medicine, Albert Einstein College of Medicine, New York, NY, USA

**Keywords:** pleiotropy, polygenic score, common variants, aging, lifespan, longevity, centenarians, proteomics

## Abstract

The highly polygenic nature of human longevity renders cross-trait pleiotropy an indispensable feature of its genetic architecture. Leveraging the genetic correlation between the aging-related traits (ARTs), we sought to model the additive variance in lifespan as a function of cumulative liability from pleiotropic segregating variants. We tracked allele frequency changes as a function of viability across different age bins and prioritized 34 variants with an immediate implication on lipid metabolism, body mass index (BMI), and cognitive performance, among other traits, revealed by PheWAS analysis in the UK Biobank. Given the highly complex and non-linear interactions between the genetic determinants of longevity, we reasoned that a composite polygenic score would approximate a substantial portion of the variance in lifespan and developed the integrated longevity genetic scores (*iLGSs*) for distinguishing exceptional survival. We showed that coefficients derived from our ensemble model could potentially reveal an interesting pattern of genomic pleiotropy specific to lifespan. We assessed the predictive performance of our model for distinguishing the enrichment of exceptional longevity among long-lived individuals in two replication cohorts and showed that the median lifespan in the highest decile of our composite prognostic index is up to 4.8 years longer. Finally, using the proteomic correlates of *iLGS*, we identified protein markers associated with exceptional longevity irrespective of chronological age and prioritized drugs with repurposing potentials for gerotherapeutics. Together, our approach demonstrates a promising framework for polygenic modeling of additive liability conferred by ARTs in defining exceptional longevity and assisting the identification of individuals at higher risk of mortality for targeted lifestyle modifications earlier in life. Furthermore, the proteomic signature associated with *iLGS* highlights the functional pathway upstream of the PI3K-Akt that can be effectively targeted to slow down aging and extend lifespan.

## INTRODUCTION

The human lifespan is a complex trait that reflects the interplay of numerous socioeconomic factors and genetic predispositions. Its narrow-sense heritability (*h*^2^) has been estimated in the range of 15 to 33%^1,2^. Recently, a lower estimate (<10%) has been reported after correcting for assortative mating^3^. The magnitude of missing heritability suggests that a large portion of heritable variation in human survival may come from small effects of numerous loci spread widely across the entire genome^4^. In fact, findings from moderately powered genome-wide association analysis (GWASs)^5–7^ are consistent with the perceived polygenic architecture of human lifespan, healthspan, and longevity.

The extensive polygenicity underlying human lifespan has some immediate implications. With a finite number of genes and a theoretically infinite number of age-related traits and endophenotypes, extensive pleiotropy would be inevitable^8,9^. The shared genetic component across the correlated traits tags the common mechanisms amenable for drug development to slow the aging process and increase the healthspan^10,11^. If only a handful of rare alleles would underlie exceptional longevity (here, longevity perceived as a fitness module), then these alleles were expected to rapidly ascend to high frequency and continuously shift the population mean life expectancy to a higher optimum. This is in contrast to the notion that there is, in fact, a limit to the maximum human lifespan^12^. Furthermore, if only a few rare loci underlie exceptional longevity, then the mutational target would be extremely narrow and, therefore, highly sensitive to population dynamics. In populations with a small effective size (*N*_*e*_) or appreciable founder effect, the strong impact of genetic drift would have supplanted other evolutionary forces and ultimately either drive the longevity-increasing rare variants to fixation or complete removal from the gene pool^13^. In the absence of earlier observations among bottlenecked and isolated populations who acquired such extraordinary longevity in an evolutionarily short period, the extent of missing heritability attributable to rare variants remains to be determined.

We posited an infinitesimal model for longevity in which variation in human lifespan is primarily determined by the additive effect of many segregating common variants (MAF> 1%), each with a small effect size. Given the additive model, we reasoned that the polygenic score would approximate a substantial proportion variance in lifespan among long-lived individuals. Leveraging the pervasive pleiotropy, we modeled highly complex and non-linear interactions among segregating variants to construct the integrated longevity genetic scores (*iLGSs*) for distinguishing differential survival. We studied a cohort comprising 515 Ashkenazi Jewish centenarians and 442 ethnically matched controls with known age at death (**Supplementary Figure** S**1**). We validated the predictive capacity of *iLGS* for distinguishing enrichment of exceptional longevity among the Wellderly cohort (*n* = 510) and Medical Genome Reference Bank (MRGB) cohort (*n* = 2,570) and subsequently converted the model to 3.8 million single nucleotide polymorphism (SNP) weights using the European descent portion of the Genetic Epidemiology Research on Adult Health and Aging (GERA) cohort (*n* = 62,268). Finally, we investigated proteomic correlates of exceptional longevity using *iLGS* and identified drugs with repurposing potentials for ameliorating the aging process. A schematic overview of the study is provided in **Extended Figure 1**.

## RESULTS

### Tracking the allele frequency changes as a function of viability

Selective pressure early in life favors maximization of the fetal viability and fecundity during adulthood, although later in life, it attenuates maximizing the homeostasis maintenance^14–17^. The adaptive response to changing selective pressure often entails allele frequency changes at many segregating loci influencing the trait^9^. Determinants of the evolutionary trade-off between viability and fecundity characterize the polygenic response that results in the fluctuation of allele frequencies across different age groups^18^. To identify variants underlying exceptional longevity, we adopted a regression framework similar to that of Bergman et al.^19^, which tracks allele frequency changes across different age bins. Variants with a negative impact on longevity are expected to be naturally purged from the centenarian gene pool, while ones with a positive contribution to healthy aging are expected to be enriched in the gene pools toward the extremities of the human lifespan.

In constructing our sliding regression framework (see **Materials and Methods**), we assumed that due to the specific age structure of our discovery cohort (between the ages of 56 and 111), the fitness cost of pro-aging alleles monotonously decreases their presence in the gene pool, while pro-longevity alleles will monotonously increase in frequency. Neutral alleles with no effect on longevity will appear with constant frequency during aging. Hence, the regression slope would reflect the overall direction of the effect conferred at each variant by the minor allele. We tested 8,090,427 variants in total and identified 34 independent lead SNPs at the genome-wide significance threshold (*P* < 5E-8) **(Supplementary Figure 2** and **Supplementary Table 1)**. These variants are sex independent as allelic frequencies in each age bin were corrected for the gender effect (see **Materials and Methods**). Among the 34 variants, two are coding and the rest non-coding (mapping to intronic and intergenic regions), and 20 variants tagging expression Quantitative Trait Loci (eQTL) **(Supplementary Table 2)**. As proof of principle, we also tracked the frequency of each of the three *APOE* haplotypes (ε2, ε3, and ε4) tagged by rs429358 and rs7412 across directly genotyped and phased samples. As expected, we identified a significant negative effect (*β* = -21.84, *P* = 4.25E-7) for *APOE*-ε4 haplotype **(Supplementary Figure 3)**.

Several genomic loci included multiple prioritized genes, including two of the previously reported contributors of aging, such as *LPA* and *LDLR*^6^. The strongest association signal for longevity (i.e., rs76430661) was on 5q35.3 where six genes were prioritized (*COL23A1, HNRNPAB, N4BP3, ZNF454, ZNF879*, and *PRELID1*. (**Supplementary Figure 4**). The variant physically maps to the second intron of *COL23A1* and is an eQTL for all the prioritized genes apart from *ZNF454* which is prioritized based on chromatin interactions. While none of the prioritized genes in the locus has been previously described in the context of aging, upregulation of a closely related family member of *HNRNPAB* is suggested to prevent age-dependent cognitive decline in Alzheimer’s disease (AD) mouse models^20^.

The largest two risk loci on 6q26 and 19q13 (tagged by rs41272112 and rs147053538), each with 16 and 50 genes respectively, represented the highest density of prioritized coding genes physically located in the locus (**Supplementary Table 3**). The lead SNP on 6q26 (rs41272112) maps to the exon 26 of *LPA* and the risk locus is an eQTL for 8 genes including *LPA* itself and *PLG* and showing chromatin interactions with *IGF2R, AGPAT4*, and *FNDC1* (**Supplementary Figure 5**). Exposure to high Lp(a) (lipoprotein A) levels is associated with coronary heart disease^21^ and was recently shown to be causally related to shortened parental lifespan in the UK Biobank^22^; nevertheless, the relationship between Lp(a) concentrations and all-cause mortality among patients with established cardiovascular diagnosis is debated^23^. Given that the prioritized missense variant in the locus is not damaging (CADD: 0.06, PrimateAI: 0.30), it is likely that its negative association with longevity (*β* = -0.21, *P* = 4.2E-08) is mediated through a more stable Lp(a) resulting from the c.4262G>A mutation. It is worth noting that the risk locus also shows chromatin interactions with the insulin-like growth factor 2 receptor (*IGF2R*), which has been identified to be associated with parental longevity^7^. Furthermore, coding variant rs3798220, which is in complete LD with the prioritized tag SNP, has been identified to show the most substantial individual-level effect in “lost healthy life years” across the participants of the UK Biobank and FinnGen cohort^24^.

The lead SNP rs147053538 on 19q13 maps to the intergenic region between *PSG11* and *PSG7*, and the risk locus is an eQTL for six genes including *ARHGEF1* and shows chromatin interactions with 13 additional genes including *CEACAM1* (**Supplementary Figure 6**). Rho Guanine Nucleotide Exchange Factor 1 (*ARHGEF1*) was identified to be crucial in angiotensin II-induced hypertension, and its inactivation has been suggested as an amenable target for the treatment of high blood pressure^25^. Given the proposed association of hypertension with late-stage dementia and Alzheimer’s disease^26,27^, the net effect of the risk loci in modulating the expression of *ARHGEF1* merits further investigation. Remarkably, age-dependent upregulation of carcinoembryonic antigen-related cell adhesion molecule 1 (*CEACAM1*) is identified to result in endothelial impairments and promotion of atherosclerotic plaque formation during aging^28^. Whether the favorable impact of the locus on longevity is mediated through the downregulation of *ARHGEF1* expression or *CEACAM1* (or both) is not clear, but we speculate that the locus implicates an important cardiovascular component of longevity that can be therapeutically targeted for ameliorating aging.

The most prolific regulatory activity was observed by rs11556579, where 43 genes were prioritized by both eQTL mapping and chromatin interactions, including *MAN1B1* on 9q34 (**Supplementary Figure 7**). ER mannosidase I (Man1b1) is involved in the intracellular clearance of misfolded alpha1-antitrypsin^29^. Accumulation of misfolded α1-antitrypsin plaques in the lesions of AD is previously reported^30^, and it has been shown that properly folded protein protects against amyloid-β-induced toxicity in microglial cells^31^. Moreover, treatment of type-2 diabetes mouse models with α1-antitrypsin rescues glucose intolerance and normalizes blood glucose levels^32^. In light of recent findings on the significance of heme homeostasis in longevity and the proposed mechanism of action for metformin^33^ that mimics α1-antitrypsin protection against heme oxidation, it is highly likely that the favorable impact of the locus is mediated through the regulation of *MAN1B1* expression that ensures conformational acuity of α1-antitrypsin. The expression of the gene is downregulated in AD^34^, and we speculate that the risk locus implicates the link between T2D and AD.

Additionally, the lead SNPs on 2q24 and 2q32 (i.e., rs6757605 and rs59642822, respectively) tagged eQTL loci that regulate the expression of *GPD2* and *GLS1* respectively (**Supplementary Figure 8**). Suppression of *gpd-2* in C. elegans is shown to further extend the life span of *daf-2* mutants^35^ and inhibition of *GLS1* ameliorated age-related pathologies by eliminating senescent cells in aged mice^36^. Given that downregulation of both genes results in extended lifespan in animal models, it is likely that the favorable effect of these two loci on human longevity is mediated by attenuation of gene expression by eQTLs in LD with the lead SNPs.

Overall, positional, eQTL, and chromatin interaction mapping prioritized 332 genes of which 191 genes are exclusively the target of expression alteration by the 20 eQTLs. Gene-set analysis suggests a shared biological function that is enriched in the CD40 pathway, thyroid hormone-mediated pathway, osteoblast proliferation, and apolipoprotein binding, however, significance levels do not withstand the multiple-test correction (**Supplementary Table 4**).

### *iLGS* model construction

Multimorbidity is a common phenomenon during aging^37^. The shared genetic component among age-related traits (ARTs) and age-related diseases (ARDs) results in cross-trait pleiotropy and underlie certain multimorbidities^38^. Given the limited size of our discovery cohort, any polygenic derivation of additivity of effect solely based on the few associated signals (from our sliding regression framework) is massively underpowered and not capable of capturing the full spectrum of additive variance underlying longevity. However, given the shared pathways among ARTs and ARDs^39^, we reasoned that combining the polygenic effect among age-related pleiotropic traits enables the robust approximation of the additive liability from variants in the common pathways related to human lifespan.

To explore the pleiotropic landscape of the 34 variants identified in the sliding-regression framework, we carried out a phenome-wide association study (PheWAS) using GWAS summary statistics primarily from the UK Biobank (see **Materials and Methods**). Overall, we tested the association of 1,504 traits across 28 domains, of which 223 unique traits and diseases across 17 domains were identified to be significantly associated with the candidate variants (**Supplementary Table 5**). The association signals were significantly enriched across three domains including Skeletal (*P* = 1.0E-05, OR = 4.39), Metabolic (*P* = 6.8E-05, OR = 1.90), and Cognitive domains (*P* = 6.3E-05, OR = 3.20) (**Supplementary Table 6**). The most significant associations across these three domains included traits such as heel bone mineral density (skeletal domain), impedance measures of body fat percentages (metabolic), and overall cognitive performance.

Given the ubiquitous pleiotropy revealed in the PheWAS analysis, we constructed a model that integrates the complex interplay of pleiotropic ARTs in shaping longevity. Here, we consider genes as the units determining longevity and presume that total contribution to longevity is better approximated by the additive effect of their constituent functional variants on pleiotropic traits determining the evolutionary trade-offs throughout the life history of the species^40^. Traits identified by PheWAS analysis either causally influence longevity or simply arise from the spurious pleiotropy due to linkage disequilibrium (LD)^41^. Regardless of the true nature of the genetic correlation between these traits and longevity, we reasoned that a model summarizing the total additive genetic variance conferred by genetically correlated traits while accounting for multicollinearity among them would yield a descriptive statistic that can be used to distinguish survival.

We applied a stacked ensemble method to construct the **integrated longevity genetic score** (*iLGS*). We first randomly split the Einstein LonGenity cohort into the derivation (*n* = 715) and validation (*n* = 237) sets. We then used polygenic risk scores of 53 select PheWAS-identified traits and 34 additional UK Biobank blood and urine biomarker traits (See **Materials and Methods**). We trained the ensemble model on 65% of the derivation dataset and tested it on the remaining 35%. Upon five-fold cross-validation, our model achieved an area under the curve (AUC) of 0.87. A schematic overview of the model construction is provided in **Extended Figure 2**. The final stacked Elastic-net regression framework shrank the coefficients of ten traits to zero, and the final score was constructed using the coefficients of the remaining 72 traits and 297 interaction terms (See **Materials and Methods**). The size of adjusted coefficients in the final model is provided in **Supplementary Tables 7 & 8**. As expected, the pattern of genetic correlation *(r*_*g*_) among the traits included in our model revealed several distinct clusters (**Figure 1**): (1) The biggest cluster includes different body impedance measures that are significantly correlated with different metabolic and cardiovascular traits (including systolic blood pressure, diabetes, birth weight, and the birth weight of first child), different metabolic traits and anxiety; Impedance measures are negatively correlated with numerous reproductive traits, which include age at menarche, age at first sexual intercourse, age at first birth and age at menopause; Fat impedance measures are also negatively correlated with different measures of fluid intelligence and usual walking pace; (4) Among the biomarker traits the greatest number of intra-domain correlations belong to C-reactive protein and triglyceride (each with 28 significant inter-domain genetic correlation), which accounts for most intra-domain correlations.

**Figure 1:**
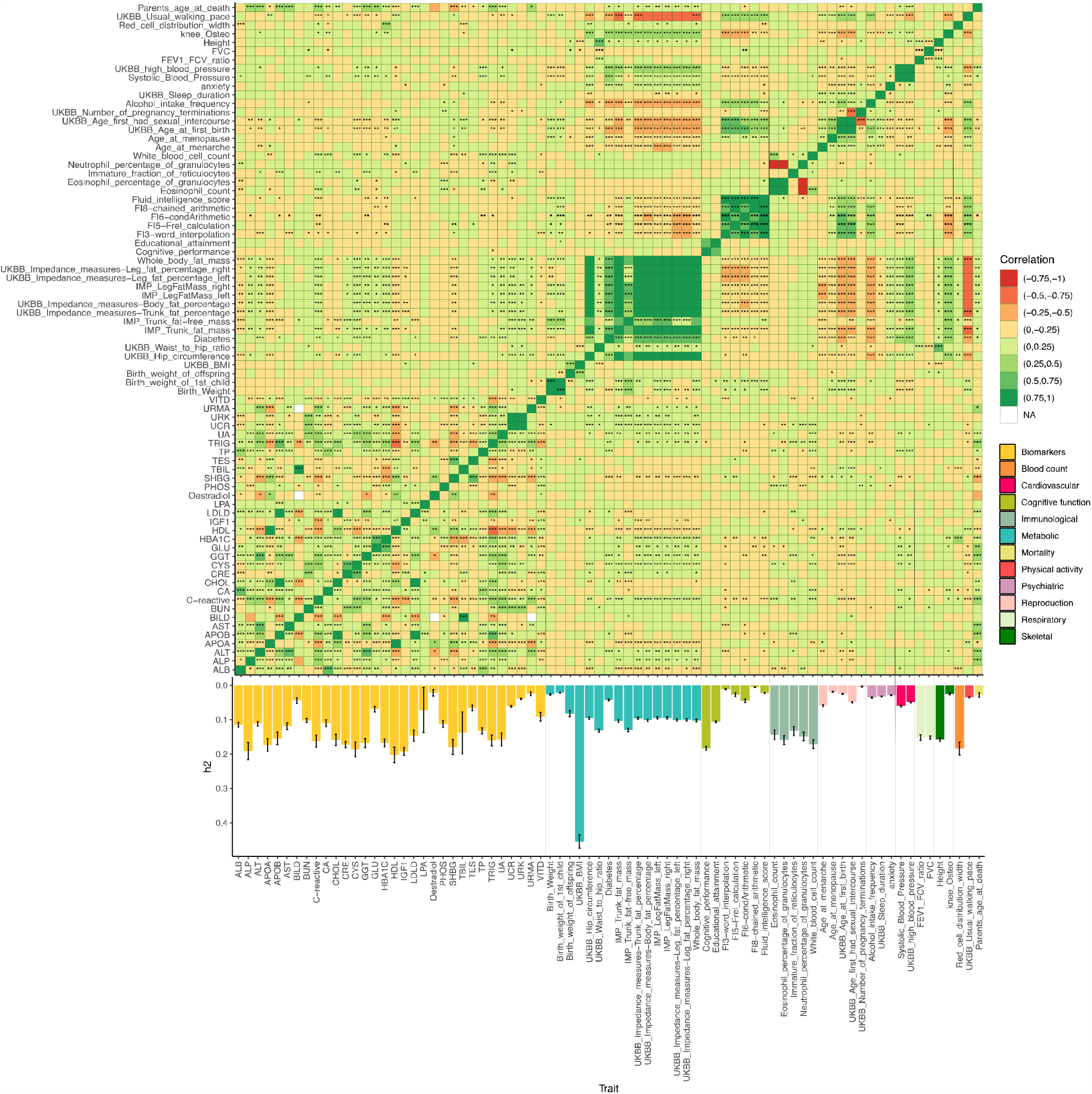
Heatmap plot showing the pattern of genetic correlation and the magnitude of narrow-sense heritability across the 78 traits used for constructing the iLGS. Pairwise genetic correlation (*r*_*g*_, n= 3,003) across the 78 traits used in constructing iLGS was calculated using bivariate LD score regression. Positive and negative genetic correlations are shown in green and red respectively. P-values for the significance of genetic correlations are FDR corrected using the Benjamini-Hochberg method. The magnitude of the *r*_*g*_ significance is indicated by the asterisks. The bars at the bottom of the plot show the magnitude of narrow-sense heritability across the 78 traits. The colour of each bar corresponds to the respective domain of the trait; (ALB: Albumin, ALP: Alkaline phosphatase, ALT: Alanine aminotransferase, APOA: Apolipoprotein A, APOB: Apolipoprotein B, AST: Aspartate aminotransferase, BILD: Direct bilirubin, BUN: Urea, CA: Calcium, CHOL: Cholesterol, CRE: Creatinine (in serum), CYS: Cystatin C, GGT: Gamma glutamyltransferase, GLU: Glucose, HBA1C: Glycated haemoglobin, HDL: HDL cholesterol, IGF1: Insulin growth factor-1, LDLD: LDL cholesterol, LPA: Lipoprotein A, PHOS: Phosphate, SHBG: Sex hormone binding globulin, TBIL: Total bilirubin, TES: Testosterone, TP: Total protein, TRIG: Triglycerides, UA: Urate, UCR: Creatinine in urine (enzymatic), URK: Potassium in urine, URMA: Microalbumin in urine, VITD: Vitamin D, FVC: Forced vital capacity, FEV1: Volume that has been exhaled at the end of the first second of forced expiration).

Across the individual PGS (polygenic score) terms, high-density lipoprotein (HDL) cholesterol, fluid intelligence (chained arithmetic measure), and forced vital capacity are positive top contributors to longevity, while glycated hemoglobin (HbA1c), apolipoprotein B, and body mass index (BMI) are top negative contributors to longevity. Our model also entails the interactions among traits (**Extended Figure 3** and **Supplementary Figure 9)**. Since the *iLGS* is linearly correlated with longevity, traits with multiple interacting partners can potentially tag canonical longevity features. Several interesting insights are immediately discernible from the interacting pairs in our model. Across the 12 domains, traits in the “biomarkers” category have the most interacting partners. Out of the 35 blood and urine biomarkers included in the model, 32 interact with at least three other traits. The highest absolute number of interactions is mediated through the insulin-like growth factor 1 (IGF1) (17 interacting partners), which is followed by the rheumatoid factor (*n* = 15), alkaline phosphatase, and estradiol (both with 14 interacting partners) **(Extended Figure 3)**. Among the remaining domains, traits within the “cognitive function” class have the second-highest number of interactions with other traits. In this domain, educational attainment interacts with 14 other traits, representing the most important non-biomarker traits in describing the additive variance in our model **(Extended Figure 3)**. Traits in the metabolic, immunological, and reproduction domains, each with 45, 34, and 29 total interactions, respectively, account for most of the remaining cross-trait interactions in our model. When traits were considered individually, age at first birth, anxiety, and walking pace has the most interacting partners, following educational attainment **(Extended Figure 3)**.

Based on our model, the interaction between fluid intelligence and serum total bilirubin has the most favorable contribution to longevity, and the interaction between IGF1 and direct bilirubin has the most negative effect on longevity. Across the individual PGS terms, high-density lipoprotein (HDL) cholesterol, fluid intelligence (chained arithmetic measure), and forced vital capacity were revealed as the top contributors to increased longevity, and Glycated hemoglobin (HbA1c), Apolipoprotein B, and body mass index (BMI) were identified as the top negative contributors to longevity.

### *iLGS* distinguishes differential survival

Using the coefficients derived from our stacked model, we computed the *iLGS* across 59,534 individuals with European ancestry in the GERA cohort and converted the scores to a set of 3.8 million variant weights (See **Materials and Methods** and **Supplementary Figure 10**). We subsequently assessed the risk prediction performance of *iLGS* for distinguishing differential longevity in three independent cohorts.

Using the validation portion of our in-house centenarian cohort (*n* = 238), we applied a multivariate Cox proportional hazard model to evaluate the association of *iLGS* with age at death while controlling for sex, *APOE4* status, and their interaction with *iLGS* (See **Materials and Methods**). *iLGS* was associated with delayed age at death with a hazard ratio (HR) of 0.09 (95% CI = [0.05, 0.18], *P* = 6.9E-13) per standard deviation of *iLGS* (**Figure 2a**). This is equivalent to a 3% decrease in the baseline mortality hazard per one unit increase in the *iLGS*. Sex is a well-known factor involving in exceptional longevity^42^ and, expectedly, we identified a significantly increased hazard rate for mortality among males against the baseline hazard (**Figure 2a**; HR = 1.84, 95% CI = [1.25, 2.73], likelihood ratio test in Cox proportional hazard model, *P* = 2.10E-3). However, no statistical interaction between *iLGS* and sex was identified for mortality hazard (**Figure 2a**; HR = 0.43, 95% CI = [0.17, 1.10], *P* = 7.76E-2), which indicates that the association of *iLGS* with survival is largely sex-independent and cross-gender difference in overall survival is not influenced by any sex-specific effects of *iLGS*. Furthermore, given the well-known effect of *APOE* haplotypes on human longevity^43,44^, we also investigated the association of all these haplotypes with age at death and modeled the interaction of *APOE*-ε4ε4 genotype with the *iLGS* in our multivariate Cox regression model (See **Materials & Methods**). Neither of the *APOE* haplotypes was significantly associated with mortality in our validation set (**Figure 2a)**. Similarly, the effect of *iLGS* on survival appears to be independent of *APOE*-ε4 status as we did not identify any evidence of statistical interaction between *iLGS* and the *APOE*-ε4ε4 haplotype (HR = 0.53, 95% CI = [0.12, 2.24], *P* = 3.87E-1) **(Figure 2a)**. We carried out a stratified Kaplan-Meier analysis across the quintiles of *iLGS* to investigate if survival curves are significantly different across the top and bottom quintiles compared to the interquintile range (IQR) **(Figure 2b** and **Extended Figure 4)**. The cumulative incidence of death was significantly different between the top quintile 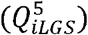 and IQR (log-rank test, *P* = 247E-5) and between the bottom quintile 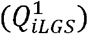 and IQR (*P* = 2.75E-27). A comparison of Kaplan-Meier functions across deciles revealed that, on average, individuals with *iLGS* in the top decile live up to 4.8 years longer. Furthermore, the risk of death for individuals in the bottom two quintiles (i.e., bottom 40%) is maximum before the age of 85, while for individuals in the top quintiles, the risk of death does not peak until over the age of 95 years (**Extended Figure 4**). The better survival outcome across the top quintile of *iLGS* is consistent with the coefficient estimates from the Cox proportional hazards model assessing HRs across the quintiles of *iLGS* (**Figure 2c**).

**Figure 2:**
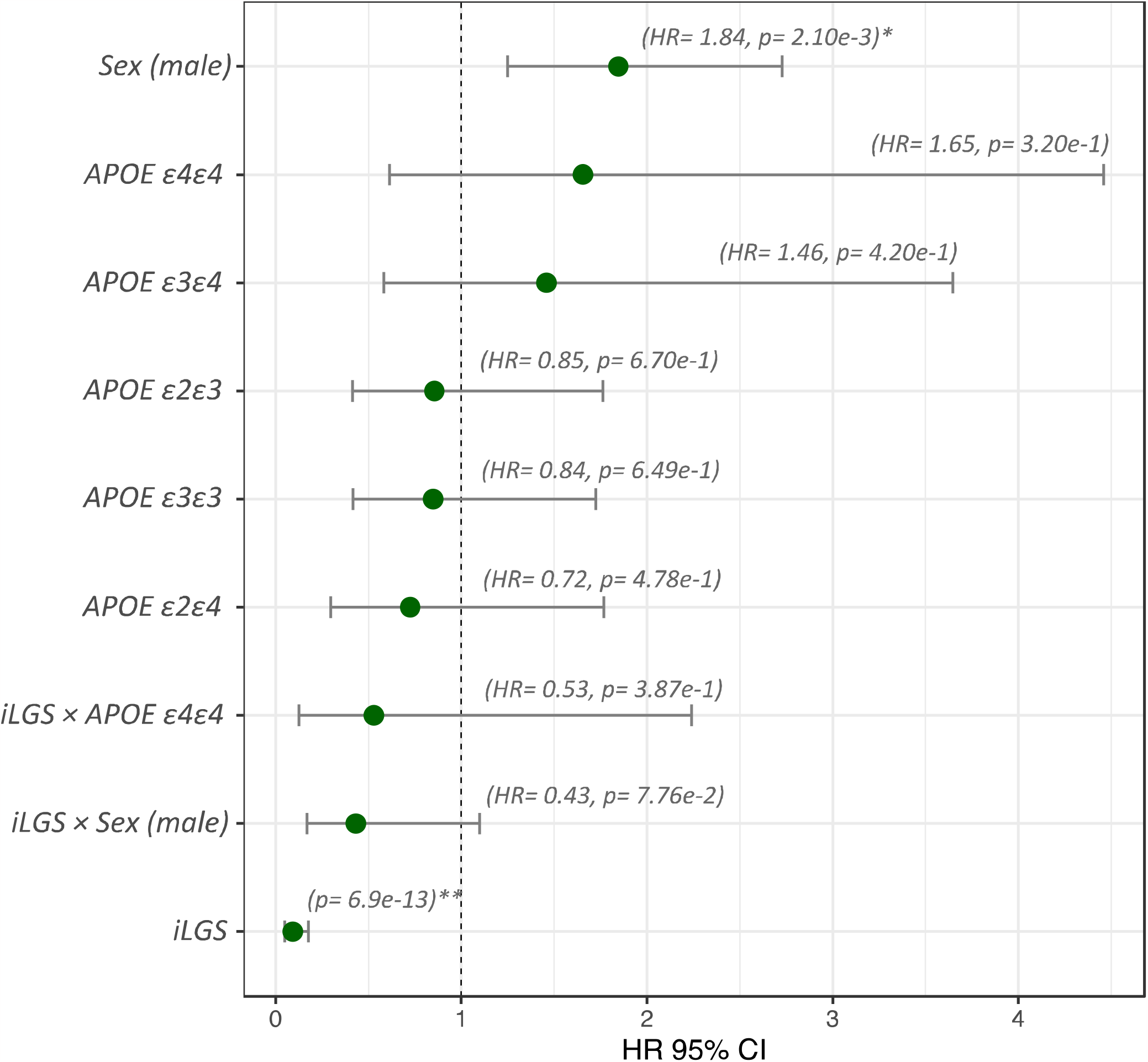
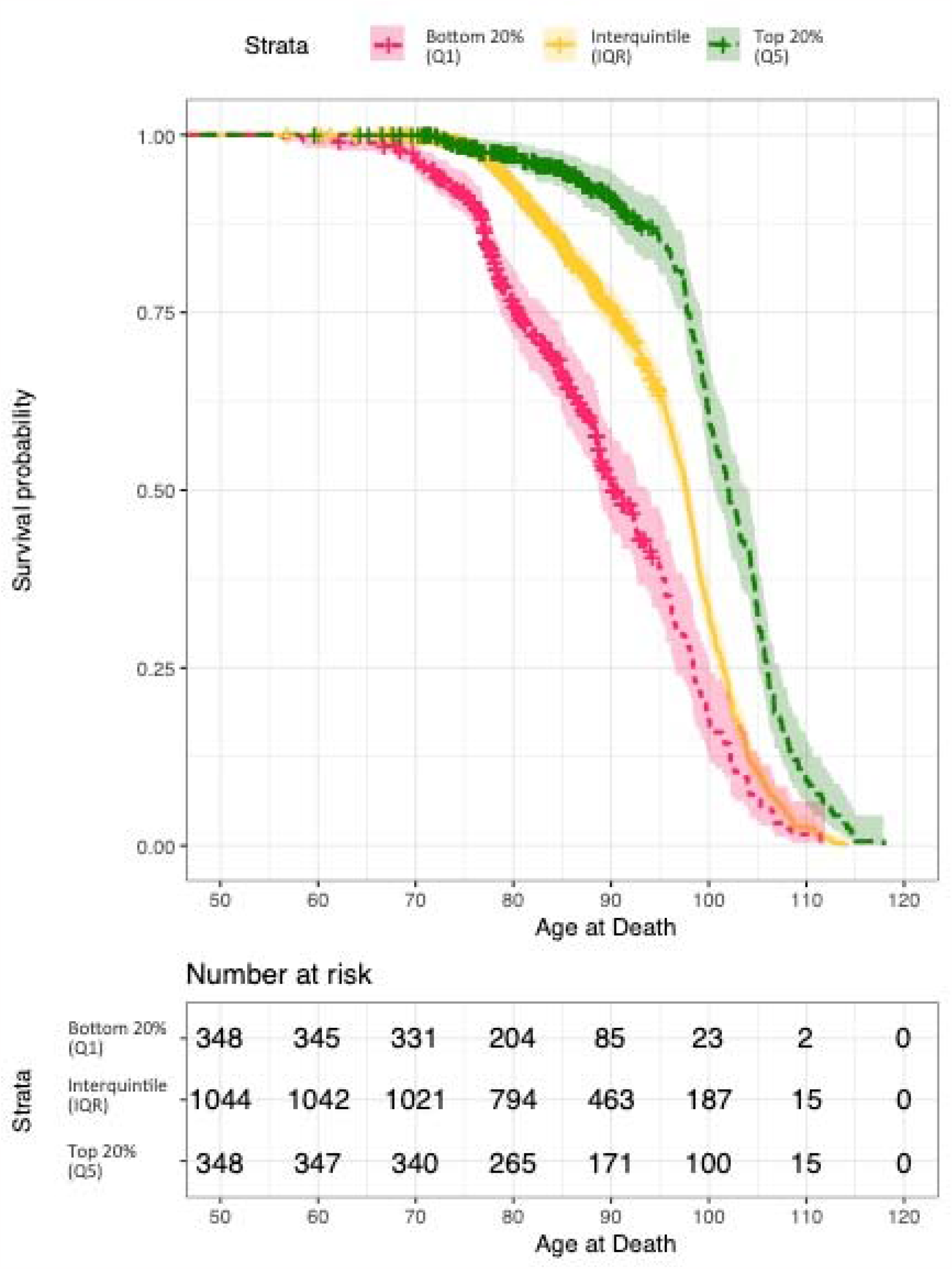
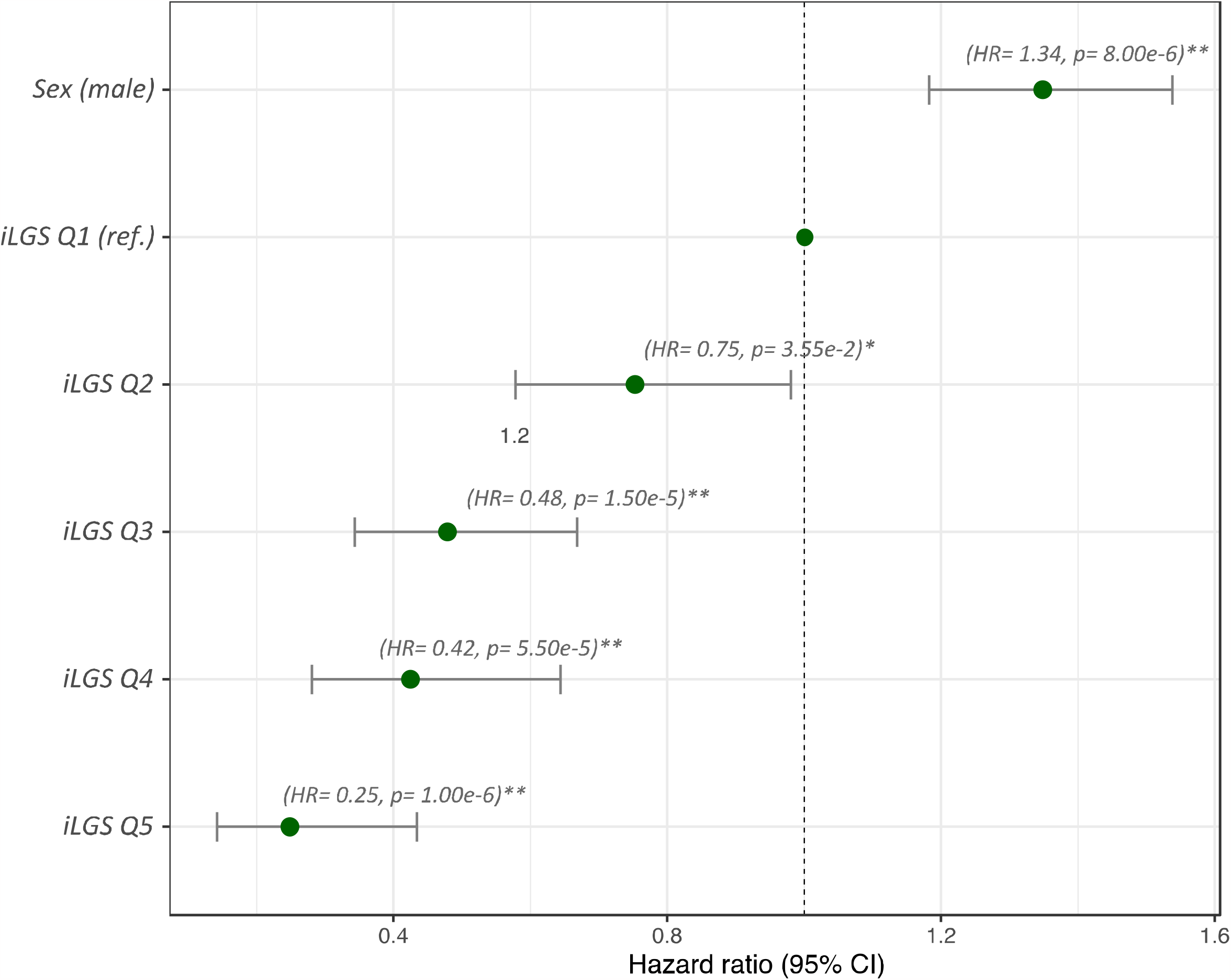
The association of iLGS with longevity and differential survival across the quintiles of iLGS. a. Cox regression hazard ratios (HR) for the association of gender, *APOE* haplotypes and iLGS with age at death. An HR greater than 1 suggests an increased risk of death, and an HR below 1 indicates a smaller risk. Expectedly, the risk of earlier death is increased among males. Neither of the *APOE* haplotypes was revealed to be significantly associated with age at death in the study population. However, the direction of HRs was consistent with the unfavourable effect of *APOE* E4 haplotypes on longevity. We also modelled the interaction of iLGS with *APOE* E4 haplotype and gender to confirm that the association is independent of the gender effect of *APOE* haplotype status. b. Stratified Kaplan-Meier curves showing differential survival across the top 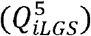 and bottom quintile of iLGS 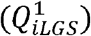 in comparison to the survival of individuals with iLGS in the interquintile range (log-rank test between the top quintile 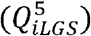 and IQR: p=2.47e-5, log-rank test between the bottom quintile 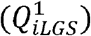 and IQR: p= 2.75e-27) c. Plot depicting the Cox regression HRs for the association of iLGS quintile with age at death. Given that our inhouse cohort is primarily ascertained to include centenarians, we adopted the 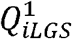as the baseline hazard and calculated the HRs for the remaining quintile against the hazards of 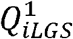 . We also included gender as a covariate to adjust for the sex effect.

We replicated the association of the top and bottom *iLGS* quintiles with three age categories (age at last contact: `≥ 95, [90, 94], and <90) in two independent cohorts, which include the Scripps Wellderly cohort`^45^ and the Medical Genome Reference Bank (MRGB) cohorts, two independent cohorts specifically ascertained to comprise a healthy aging population. However, since the absolute “age at death” for participants of these two cohorts are not available, we formulated the null hypothesis ( *H*_0_) as no association between the top 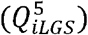 and bottom 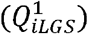 quintiles with age category at the last point of contact, and tested for evidence against *H*_0_ using a Fisher’s Exact test. In the Wellderly cohort, the top quintile 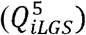 was significantly associated with both the`≥95 and [90, 94] age categories` (**Figure 3a**). Individuals in 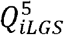 are 3.67 times more likely to surpass 95 years of age (95% CI = [1.68, 7.95], *P* = 6.68E-4) and 2.49 times more likely to fall into the [90, 94] age bin (95% CI = [1.70, 3.73], *P* = 6.89E-6) than the rest of the *iLGS* strata. Here, the definitive lifespan is not known, and “age at last contact” is used as an approximate proxy for lifespan. As such, age categories further away from the two oldest age bins (i.e., ≥95 and [90, 94]) are necessarily comprised of individuals who have already achieved their maximum lifespan and individuals who will live longer and proceed to higher age bins. Therefore, the odds of individuals with favorable polygenic background (i.e., 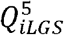 is expected to be attenuated in the lower age bins and continue to increase as we move to upper age-bins. This is consistent with the observed 1.47-fold increase in the odds of association between the 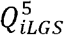 and the `≥95 age category (compared to the [90, 94] age bin). Since individuals of the Wellderly cohort are essentially devoid of age-related conditions, the lack of association between the bottom quintile` 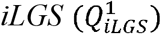 and age categories is not surprising (**Figure 3a**). In the MRGB cohort, 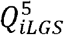 is significantly associated with both the ≥95 and [90, 94] age bins (**Figure 3b**). Here, individuals in 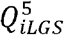 are 4.27 times more likely to surpass 95 years of age (95% CI = [3.43, 5.33], *P* = 7.88E-47) and 3.66 times more likely (95% CI = [2.96, 4.53], *P* = 2.37E-30) to reach the [90, 94] age category than the rest of the population. Individuals with *iLGS* in the bottom quintile 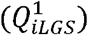 were significantly depleted from the [90, 94] age bin (OR: 0.50, 95% CI = [0.25, 0.90], *P* = 3.02E-2). Conversely, 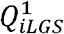 was significantly enriched among individuals in the <90 age bin (OR: 1.38, 95% CI = [1.08, 1.76], *P* =9.79E-3).

**Figure 3:**
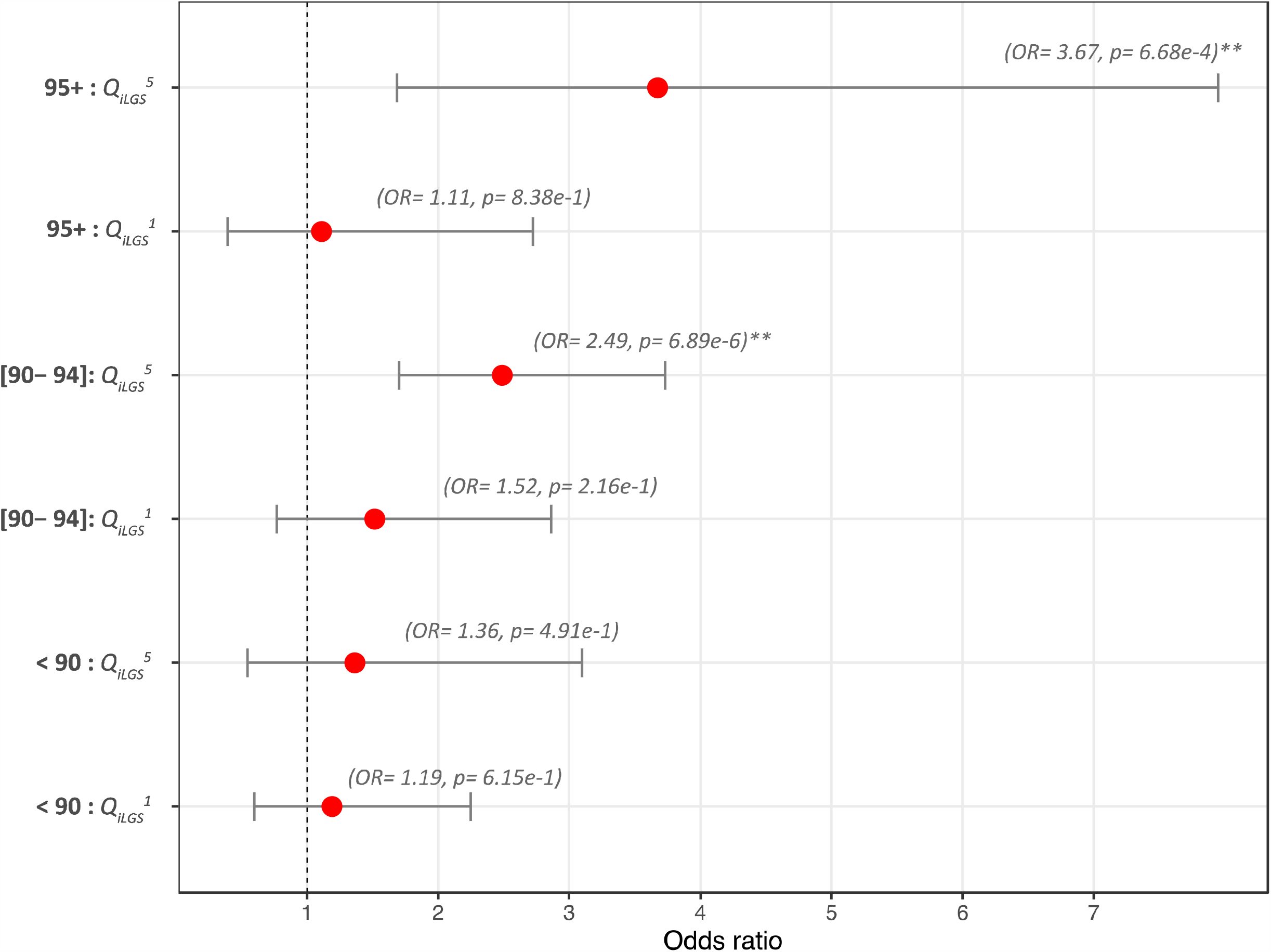
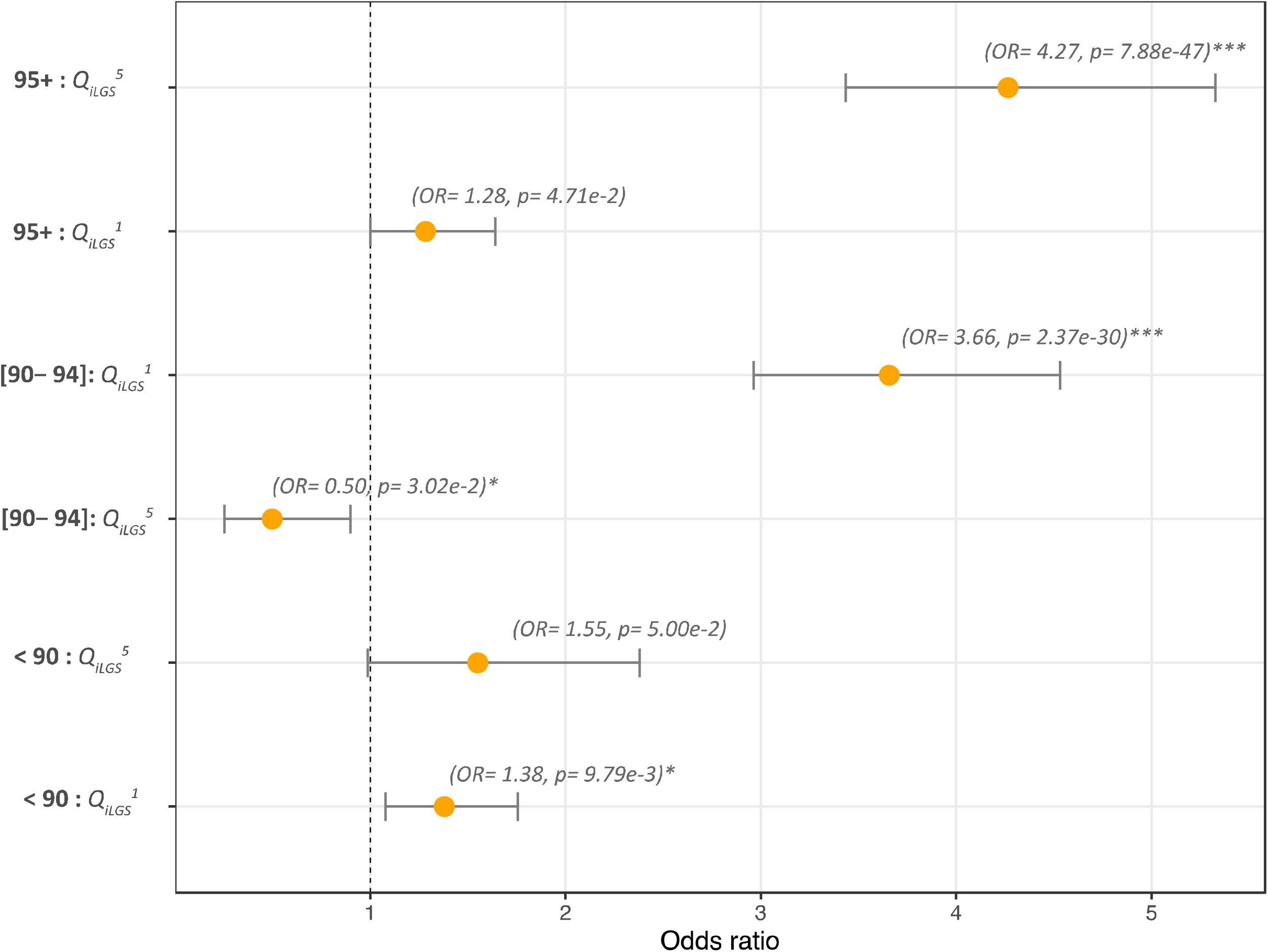
Replication of iLGS association with age at last contact in the Wellderly and MRGB cohort. a. Plot illustrating enrichment of top and bottom quintile iLGS scores (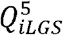 & 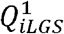 ) across three age bins (age at last contact: ≥ 95, [90-94] and <90) of the Wellderly cohort. (asterisks identify significant associations). b. Enrichment of top and bottom quintile iLGS scores (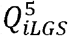 & 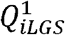 ) across three age bins (age at last contact: ≥ 95, [90-94] and <90)) of the MRGB cohort. (asterisks identify significant associations).

In both replication cohorts, the strength of association between 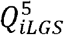 and the older age categories is significantly stronger. Given that age at death in these cohorts is undetermined, we believe the stronger signal toward exceptional longevity arises from the more homogenous cohort construct among individuals achieving the upper limits of lifespan. Using the available clinical data in the MRGB cohort, we further investigated the association between 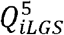 and three age-related traits: extreme obesity (BMI> 40 kg/m ), treatment history for high blood pressure and treatment history for high cholesterol. Despite the expected direction of effect across the three traits (**Supplementary Figure 11**), the protective effect of 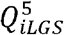 is only statistically significant against treatment for high blood pressure (OR: 0.64, 05% CI = [0.98, 0.42], *P* = 3.82E-2).

### Burden of rare variants across 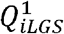 **and** 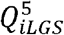

Given the association of 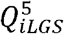 `with an extended lifespan, we compared the burden of rare pathogenic variants (PrimateAI score ≥ 0.9 and alternative allele frequency < 1%) among the top versus bottom quintile` *iLGS* carriers across centenarians and non-centenarians in our longevity cohort. A significant difference in the number of rare pathogenic variants between the 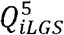 and 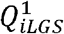 centenarians was observed (**Supplementary Figure 12**). It appears centenarians in the top quintile *iLGS* collectively carry a higher burden of rare pathogenic variants compared to centenarian carriers of 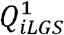 . This perhaps reflects the buffering effect of the favorable polygenic background that offset the higher burden of rare pathogenic variants among 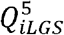 carriers. This observation is consistent with a liability threshold model, where the accumulation of liabilities from pathogenic variants is favorably offset by the higher threshold rendered by the polygenic background^46^.

### Proteomics correlates of *iLGS* and anti-aging drug repurposing

The quantitative size of *iLGS* is determined at birth and remains constant over an individual’s life course. Given the association of *iLGS* with longevity, we searched for the proteomic correlates of the score among the 237 individuals (123 males and 115 females) in our validation set. In doing so, we reasoned that proteomic predictors of aging and longevity could be detected via the patterns of association between the proteome and the *iLGS*. Briefly, we tested the association between 4,265 proteomic markers and *iLGS* among males and females separately and identified significant associations with FDR < 0.05 (See **Materials and Methods**). Among females, 37 proteins were positively associated with *iLGS*, while three – MAT2B, CLEC2B, and CMPK1 – show negative association (**Figure 4a**). Among these 40 proteins, 30 were associated with chronological age among female Ashkenazi Jews^47^ (**Supplementary Table 9**), and ten were differentially expressed among centenarians with a consistent direction of effect^48^ (**Supplementary Table 9**). The most significant positive association signals came from semaphorin 4B (SEMA4B, β = 4.94, *P* = 1.49E-6) and Pleiotrophin (PTN, β = 3.98, *P* = 1.49E-6) (**Figure 4a**). Previous studies have repeatedly replicated the association of pleiotrophin with both chronological age and extended lifespan^48–50^ and identified the association between the expression of *SEMA4B* and favorable proteomic signature of intermittent fasting^51^. On the other hand, methionine adenosyltransferase 2B (MAT2B) showed the most significant negative association with *iLGS* (β = -3.56, *P* = 2.05E-5) (**Figure 4a**). The connection of MAT2B to aging has not been studied, although its overexpression has been attributed to poor prognosis of triple-negative breast cancer^52^ and critical illness in COVID-19^53^.

**Figure 4:**
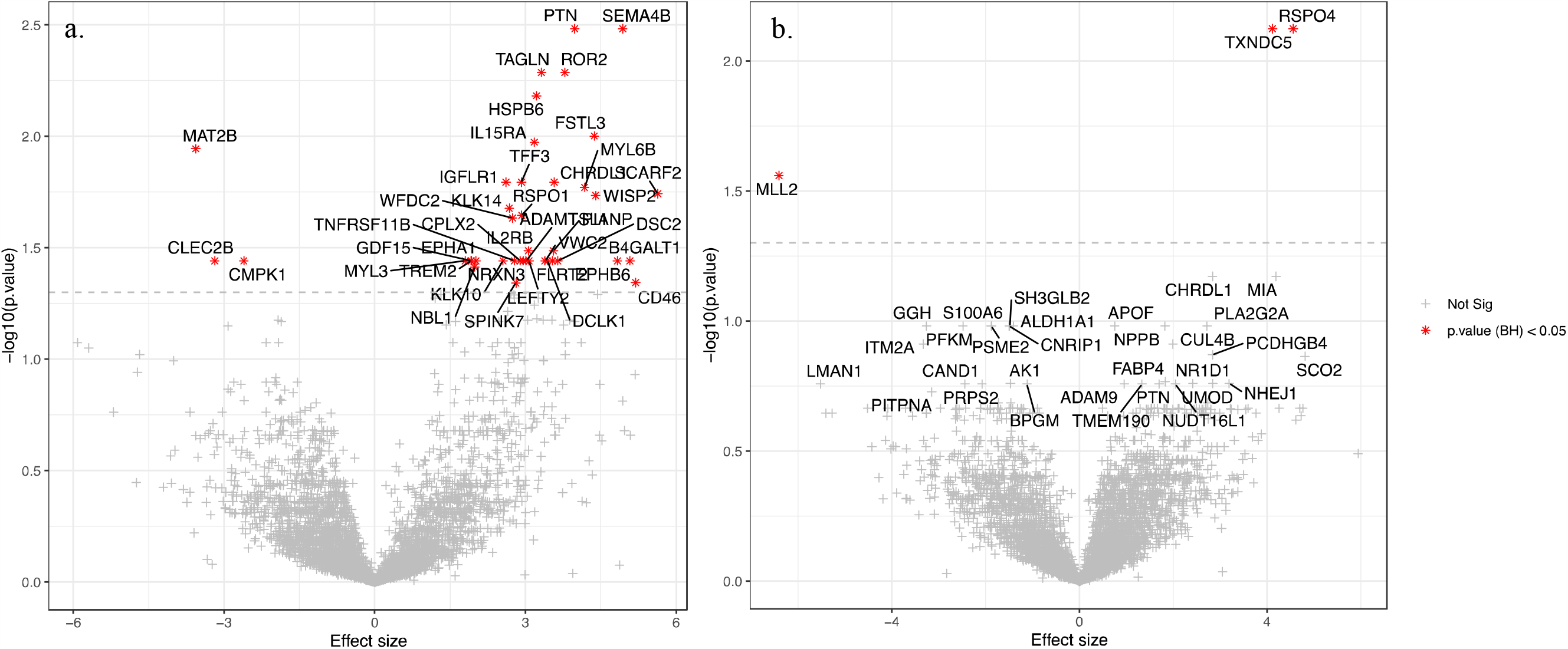
Proteomic correlates of iLGS. Volcano plot showing the association of serum proteins with iLGS among female (a) and male (b) subsets of the validation set (n= 115 and 123 respectively). The x-axis denotes the “effect size”, and the y-axis shows -log10 Benjamini–Hochberg corrected p-value of association. Proteins clustered to the right are positively correlated with iLGS and proteins clustered to the left are negatively associated with iLGS. Proteins surpassing the significance threshold at BH< 0.05 are identified by red asterisks.

Among male subjects (*n* = 123), only three proteins were significantly associated with *iLGS* (**Figure 4b**). R-spondin 4 (RSPO4) showed a positive association signal (β = 4.82, *P* = 1.85E-6), which was the most significant. This is consistent with an increased expression of *RSPO4* among centenarians as previously reported^48^. We detected a positive association for Thioredoxin domain-containing protein 5 (TXNDC5, β = 4.25, *P* = 3.28E-6). Although the association of endoplasmic reticulum (ER) protein TXNDC5 with human longevity has not been previously reported, its transcriptional upregulation has been shown to be associated with age in long-lived red sea urchins^54^. We also identified a negative association for lysine methyltransferase 2D (MLL2, β = -6.38, *P* = 4.19E-5). Among the 43 proteins significantly associated with *iLGS* in either males or females, the direction of effect was inconsistent between the genders for only three proteins: MAT2B, CELEC2B, and CMPK1 (although not significant in males) (**Supplementary Table 9**). This inconsistency might indicate the sex differences in the proteomic signature of aging.

We queried the DrugBank database (version 5.1.8)^55^ to identify drugs targeting or interacting with the 43 *iLGS*-associated proteins. We only considered drugs with ongoing clinical trial status (see **Materials & Methods**). Our rationale was that the underlying mechanism of action for drugs interacting with *iLGS*-associated proteins might implicate the pathways relevant to longevity and they could be repurposed as anti-aging treatments. We identified 25 unique drugs interacting with 11 proteins (**Table 1**). One drug, fostamatinib, interacting with at least four longevity-promoting proteins, is a selective inhibitor of spleen tyrosine kinase (Syk)^56^ that acts upstream of the PI3K-Akt pathway, which is upregulated during aging and whose downregulation promotes improved cell survival in neurons^57^. The suppression of Syk attenuates the PI3K-Akt signaling, which inhibits mTOR and activates FOXO, leading to the activation of proapoptotic processes and cell cycle arrest. Although its precise mechanism of action is unclear, the active metabolite in fostamatinib has been recently found as a novel senolytic agent^58^. Given the extent of its interaction with longevity-promoting proteins, this drug merits further investigation as a potential anti-aging treatment.

## DISCUSSION

In this study, we developed a predictive genetic score for lifespan based on GWAS summary statistics for a compendium of aging-related traits. We posited that the highly complex and polygenic architecture of human longevity is well approximated by an additive model that captures the cumulative liability conferred by pleiotropic traits associated with survival components. We assessed the performance of our model for distinguishing differential survival and showed, at the population level, *iLGS* predicts prolonged lifespan independent of gender or *APOE* haplotype status. We also established that the top quintile of *iLGS* is significantly enriched among individuals surpassing 95 years in the extant populations. Finally, we identified protein correlates of *iLGS* among centenarians and drugs that could be repurposed for potential anti-aging treatments.

In building our model, a few key assumptions were made. First, we assumed a highly polygenic architecture underlying human longevity. Our assumption of high polygenicity and genome-wide distribution of associated variants is supported by the proportional correlation of lifespan heritability with chromosomal length (**Supplementary Figure 13)**, which indicated that genomic loci with a non-neutral net effect on longevity are numerous and widely distributed across the genome. This is consistent with the overwhelmingly polygenic architecture inferred from a wide variety of complex traits^59,60^. Second, we posited that the deviation of an individual’s lifespan from the population mean is primarily due to the additive liability conferred by common variants and can be well approximated by an additive model. Our supposition of “additivity of effect” follows the premise of the “omnigenic model” of complex traits^61^, where a great portion of heritability is attributed to common variants with modest effect size^62^. Obviously, rare variants with sizable effects may still be relevant in the context of longevity, although such variants are primarily implicated for traits under a strong selective pressure^9,63^. Given the attenuated selective pressure during the aging^14,16^, variants with a substantially detrimental effect on survival are expected to be under a more lenient purifying selection. Consequently, their frequency is not necessarily bound to lower limits, and therefore, rare variants with large effect sizes on aging and longevity are expected to be scarce. Third, we modeled lifespan as a function of a set of traits that are directly or indirectly under selection. Of note, lifespan is the ultimate trait at the top of the fitness hierarchy and its variance is determined by the interplay of other fitness components, which include fertility, fecundity, and inclusive fitness. We used polygenic scores of nested traits (as a reasonable proxy for unmeasured trait values) to model the perceived interplay and derive a single composite genetic score for longevity. Unlike other biological age predictors (such as epigenetic clocks, telomere length, transcriptomic, and metabolic predictors) that are dependent on the actual chronological age and may vary over time, *iLGS* is stable throughout life and can be assessed from birth. Thus, the score can be incorporated with other quantitative predictors of age later in life when they become available to give a more accurate prediction of biological age.

Our results showed that *iLGS* generalizes well in predicting lifespan in the extant populations when the age at death is not yet available. Furthermore, we showed that in the Wellderly and MRGB cohorts of healthy aging, the odds of individuals with top quintile 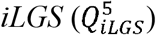 `increased sharply in the ≥95 age bin. Although we identified a protective association between` 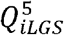 and the treatment history for high blood pressure in the MRGB cohort, we did not observe a significant association between the *iLGS* and the age at onset (or post-intervention survival) for cardiovascular complications or three cancer types (breast, prostate, and skin) in a small portion (n= 390) of our in-house cohort with available data. A number of possibilities may explain this lack of association: (1) The age distribution in our test set is relatively narrow (95 ∼ 114 years) (**Supplementary Figure 1**), and therefore, our ability to discriminate individuals based on *iLGS* strata is reduced accordingly. In particular, these complications occur in such a narrow range after age 95 that the age at onset and the survival time are indistinguishable across *iLGS* strata; (2) Heterogeneity of the investigated conditions may play an important role that is underappreciated by our model. Specifically, the underlying cause of differences in the age at onset or survival time may not be completely polygenic in nature (hence not distinguishable by our model) and simply arise due to the acquired somatic mutations in the case of cancers and lifestyle differences in the case of cardiovascular complications. This is quite an interesting possibility since studies into the congestive heart failure diagnosis have also questioned the validity of prognostic markers among centenarians^64^; and (3) Given the limited sample size, our study was underpowered.

The specific pattern of interaction between individual traits in our model shed new light on the approximate relationship between different traits on longevity. We observed that a great majority of these interactions are mediated through the insulin-like growth factor 1 (IGF1). This recapitulates the well-established significance of this hormone in human aging and underscores the relevance of interventions targeting IGF1-extended signaling network for longevity^65^. Consistent with this, we highlight the repurposing potential of fostamatinib as a senolytic drug candidate. It interacts with four *iLGS*-associated proteins (PTN, EPHA1, EPHB6, and DCLK1) and attenuates the PI3K-Akt signaling, which leads to a range of favorable outcomes that collectively promote longevity^65^. Further, we highlight the significance of our finding for proteins associated with *iLGS*. Unlike earlier studies into the proteomic correlates of longevity where it is not clear whether differential protein levels are truly causal or simply fluctuate due to aging, our results are not confounded by the chronological age.

Nevertheless, our results should be considered in light of a few limitations. First, in constructing our model, we used GWAS summary statistics from individuals of primarily European ancestry. Polygenic scores are notoriously sensitive to allele frequencies and patterns of LD that vary with ancestry^66,67^. Thus, our model will likely underperform in populations of non-European descent. Second, it remains to be determined whether the association of *iLGS* with lifespan is independent of lifestyle factors. Although we showed that the association of *iLGS* with longevity is independent of gender and *APOE* status, using the available data, we could not rule out the confounding effect of lifestyle choices. If the effect of *iLGS* on longevity is mediated through a confounder such as dietary pattern or medication intake, we expect that protein correlates of *iLGS* to be also confounded. However, we could not test for this confounding in a replication cohort with available lifestyle data like the UK Biobank dataset since summary statistics for constructing *iLGS* were primarily obtained from UK Biobank GWAS studies.

Taken together, we showed how pervasive pleiotropy could be leveraged in constructing a composite polygenetic score for longevity. The application of *iLGS* to the extant populations offers an opportunity for better risk management among individuals at the lower end of the score distribution. These individuals are more likely to develop age-related pathologies earlier in life and, therefore, benefit from lifestyle modifications. Moving forward, in expectation of more powerful GWAS studies in diverse populations, we believe our method offers a promising framework for stratifying life expectancy in the extant population.

## METHODS

### Ethics statement

In this study, we used datasets from the UK Biobank (application number 58069), as approved by the UK Biobank board, the Wellderly cohort (the Scripps Institute, La Jolla, USA) obtained from the European Genome-Phenome Archive (EGAS00001002306), the Medical Genome Reference Bank cohort obtained from the Garvan Institute of Medical Research, Australia, and the Genetic Epidemiology Research on Adult and Aging (GERA) obtained through dbGaP (request number 82588-1). The study and data access were approved by Albert Einstein Institutional Review Board, protocols 2019-9922 (reference number: 05294). All methods were carried out in accordance with the relevant guidelines and regulations, and informed consent was obtained from the research participants.

### Data sets

#### Einstein LonGenity cohort

We performed our analysis on a sub-cohort of 1,740 (722 males and 1018 females) offspring of Ashkenazi Jewish centenarians and controls recruited longitudinally in the LonGenity project at the Albert Einstein College of Medicine^68^. These individuals were genotyped on a custom array at 635,623 SNPs and subsequently underwent whole-exome sequencing (WES). Details pertaining to DNA sample preparation and sequencing are explained elsewhere^69^.

#### Wellderly cohort

For replication, we used the whole-genome sequence data from the 510 healthy elderly individuals recruited through the Scripps Institute Wellderly study. These individuals are all healthy and aged between 80 to 105 years without any chronic conditions or medication-taking history. Details pertaining to the cohort demography and the genome sequencing procedure are explained in an earlier study^45^.

#### MRGB cohort

For replication, we also used the whole-genome sequence data from 2,570 individuals (1,251 males & 1,319 females) in the Medical Genome Reference Bank (MRGB) cohort. This cohort comprises Australians who lived at least 70 years of age without any history of cancer, dementia, and cardiovascular disorders at baseline entry or study follow-ups. Gender-stratified age distributions across participants are provided in **Supplementary Figure 14**. The cohort demography and genome sequencing details are explained elsewhere^70^.

#### GERA cohort

We used a GERA (Genetic Epidemiology Research on Aging) sub-cohort of 62,268 individuals with European ancestry to convert *iLGS* to SNP weights. We used pre-computed principal component scores released by the GERA consortium to filter out 16,151 individuals with non-European ancestry.

### Quality control (QC)

We applied standard GWAS QC measures^71^ implemented in PLINK v1.90b^72^. Briefly, we removed individuals with a mismatch between the reported and inferred gender from the sex chromosomes. We restricted our analysis to unrelated individuals for all cohorts by excluding the younger participants from each pair of close relationships (up to second-degree). Our QC measures at the sample level also included removing individuals with <95% call rate, individuals with extensive runs of homozygosity, or non-European ancestry as determined by the PCA analysis. The Einstein LonGenity cohort analysis exclusively focused on the Ashkenazi Jewish cluster. At the variant level, our QC measure involved removing SNPs with < 1% minor allele frequency (MAF), SNPs with < 95% call rate among all individuals, or an extreme deviation from the Hardy-Weinberg equilibrium (HWE) at *P* `≤ 1E-6, which most likely represents poor genotyping`.

We imputed the QC’ed genotypes across filtered individuals using the Michigan Imputation server^73^ and 1000 Genomes phase 3 haplotypes as the reference panel. For the UK Biobank sub-cohort, we directly obtained version 3 of the imputed genomes from the consortium. Post-imputation QC measures across all cohorts included removing SNPs with IMPUTE2 info score < 0.5, MAF < 1%, or an extreme deviation from HWE at *P* ≤ 1E-6. For the Einstein LonGenity cohort, we merged the post-imputation genotypes with the exome data while correcting for the strand orientation. Prior to merging, genotypes from the exome data were restricted to bi-allelic loci only, and INDELs were removed. Wherever the imputed genotype was different from the exome data, we replaced the inferred alleles with the observed nucleotides from the whole-exome sequencing (WES) data.

### Principal component analysis (PCA)

To avoid stratification bias, we restricted our analysis to a single ancestry (ASJ for the Einstein LonGenity cohort and European ancestry for the replication cohorts). We used pre-imputed genotypes to calculate principal components in KING v2.2.5^74^. Before running KING, we applied a more stringent QC measure using PLINK (v.1.90b) to remove SNPs and individuals with <99% call rate. Samples were pruned to include founders only. Additionally, SNPs with MAF< 5% were removed and pruned to include uncorrelated SNPs at pairwise *r*^2^< 0.2. The top three PCAs were projected to that of 1000 Genomes phase 3 data, and samples with European ancestry were selected for downstream analysis.

### Sliding regression framework

Unrelated ASJ samples from the Einstein LonGenity cohort (*n* = 952) were divided into 48 overlapping bins according to their reported age at death. Bins were constructed to contain 30 individuals and an overlapping size of 15 individuals between the neighboring windows. The allelic frequency across 8,090,428 imputed SNPs was calculated in each bin. Details about the age distribution of participants in each bin are provided in the **Supplementary Table 10**. To remove the sex effect in the ultimate model and correct for the different male/female ratios across bins, calculated allele frequencies were regressed on to the number of females in each bin and residuals were then used in the ultimate regression model so that:

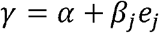

where *γ* is the age at death, *β*_*j*_ is the slope of regression, *e*_*j*_ is the sex-corrected frequency from the prior regression, and α is the intercept of regression defined by the expected mean age at death in our discovery cohort (∼70 years). We tested for an effect of gender-corrected allele frequency (*ej*) by a two-tailed t-test where we constructed our test hypothesis as below:

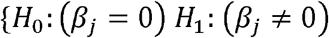

We interpreted positive *β*_*j*_ and negative *β*_*j*_ as pro-longevity and pro-aging, respectively. For each *β*_*j*_, we calculated the standard error using the formula below:

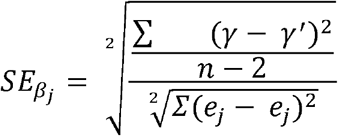

where, *γ* is the age at death, *γ*^′^ is the estimated age at death, n is the total number of bins, *e*_*j*_ is the frequency residual and *e*_*j*_ is the mean of frequency residuals. Variants surpassing the genome-wide significant threshold (*P* < 5E-8) were selected for replication analysis in the UK Biobank dataset and downstream analysis.

The number of variants in all age bins was consistent across all autosomes (**Supplementary Figure 9**).

### Functional enrichment analysis

To identify functionally relevant SNPs, we used FUMA^75^ to analyze the summary statistics from the sliding regression analysis. Variant coordinates in all analyses were defined according to the Hg19/GRCh37 assembly. We identified significant independent SNPs as variants surpassing the genome-wide significant threshold (*P* < 5E-08) and are independent of each other at *r*^2^ < 0.1. We defined LD blocks surrounding the lead SNP according to the whole-genome LD maps of the European population^76^ and included all known SNPs in the LD block (regardless of being included in the sliding regression) that are in LD with the lead SNP at *r*^2^ ≥ 0.5 for eQTL and chromatin interaction mapping. The defined risk loci, therefore, may contain SNPs that were not available in the summary statistic input from the sliding regression but are linked to the significant independent SNPs according to the 1000G reference panel. For each risk locus, we only retained SNPs with MAF ≥ 0.1 in the Ashkenazi Jewish population. Additionally, where the distance between two risk loci in neighboring LD blocks was less than 250 kb, the neighboring LD blocks were merged to a single larger risk locus. We mapped SNPs to the nearest gene within a 10 kb window for positional mapping. For eQTL and Chromatin interaction mapping, annotation was carried out over the entire risk region and genes within the risk locus, and those located outside that are linked to the SNPs in the genomic risk locus were prioritized.

### Phenome Wide Association analysis (PheWAS)

We tested the association of the shortlisted variants with 1,504 well-powered traits and disease endpoints ascertained primarily from the UK Biobank. Each SNP was interrogated for association independently, and traits surpassing the Bonferroni corrected threshold for multiple testing were reported as significantly associated. To test whether the extent of observed pleiotropy is higher than expected in specific trait domains, we defined the null distribution as an equal number of positive signals across all domains (under the null hypothesis of no differences in domain-specific pleiotropy). We used a one-sided Fisher’s exact test to compute odd ratios.

### Polygenic risk score (PRS) calculation

We calculated the additive effect of common variants on the genetic liability of traits associated with candidate variants. We used the clumping and thresholding strategy implemented in PRSice software. We computed polygenic scores for each trait independently using the formula below:

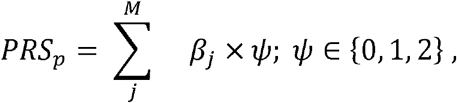

where *β*_*j*_ is the effect size (or log odds ratio (OR) for binary traits) of the allele j, *ψ* is the allele dosage for SNP j, and M is the total number of SNPs after pruning.

We followed the best practice recommendation for PRS calculation^78^. Briefly, we calculated the chip heritability (h^2^_SNP_) across each independent trait using the LD score regression^79^ and confirmed h^2^_SNP_ > 0.05. Designations of the effect allele across all summary statistics were harmonized to match those of the UK Biobank. We used PRSice-2^80^ to calculate PRSs. We implemented the standard “clumping + thresholding” method to remove highly correlated SNPs (*r*^*2*^ > 0.1) and retain only the most significant associations. Since the optimal SNP association *P*-value for inclusion in the PRS calculation is unknown a priori, we generated PRS across a range of p-value thresholds and selected the threshold providing the highest Nagelkerke R^2^.

We calculated the PRS correlation (*r*_*PRS*_) as Pearson’s correlation of standardized PRS across the 87 traits. As expected, PRS scores for traits within the same domain and related biomarkers showed a higher correlation.

### Derivation of Composite Polygenic Scores

We modeled the age at death using the scaled PRS scores across the 87 traits (53 traits from PheWAS analysis and 34 blood and urine biomarker traits). All PRSs were standardized to the unit scale (i.e., zero mean and unit standard deviation) across the entire dataset. We split our cohort into derivation (*n* = 715) and validation set (*n* = 237). To increase the statistical power of the model for predicting longevity, the derivation set was enriched for samples with age at death over 95 years.

We used a stacked model for predicting age at death based on trait PRSs. From the 87 distinct traits and endophenotypes, we first used an Elastic-net regression with five-fold cross-validation to remove non-informative PRSs. Next, the remaining PRSs were used in a polynomial regression to derive the informative interactions between the PRS scores, and finally, all interaction terms and individual PRS scores were plugged into the final elastic-net regression to remove the non-informative interaction terms. In all penalized regression steps, we tested a range of penalties to decide the optimum penalty threshold and selected the best performing model in terms of cross-validated AUC. We derived the iLGS from the formula below:

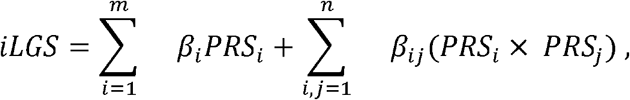

where *β*_*i*_ is the coefficient of PRS for trait *i* in the ultimate model, and *β*_*ij*_ is the non-zero coefficients for the interaction terms between PRSs *i* and *j* drawn from 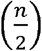, in which *n* = 77 is the total number of PRSs with non-zero coefficient from the primary elastic-net regression.

### Derivation of iLGS SNP weights

We used a subsample of 62,268 individuals with European ancestry from the GERA cohort to derive the SNP weights. We imputed genotypes across autosomes using the Michigan Imputation Server^73^ and 1000 Genomes phase 3 haplotypes as the reference panel. We excluded SNPs with IMPUTE2 info score < 0.5 and carried out standard QC on the remaining SNPs. To enhance the calibration of statistics derived from our generalized linear model, we removed loci with minor allele counts less than 20 in PLNIK (version 2.0). We calculated kinship coefficients (*φ*_*k*_) in KING (version 2.2.5)^74^ and removed related individuals up to the second degree, which is defined as *φ*_*k*_ > 0.088. Imputed and QC’ed genotypes from the remaining 59,534 individuals were used in a generalized linear regression formwork to derive *iLGS* SNP weights according to the formula below:

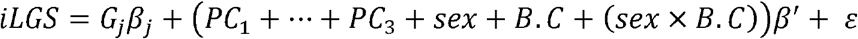

where *iLGS* is the vector of scores, *G*_*j*_ is the dosage matrix for variant j, (*PC*_1_ + ⋯ + *PC*_3_ + *sex* + *B. C* + (*sex* × *B. C*)) is the fixed-covariate matrix correcting for the top 10 principal components, sex, birth cohort (B.C), and the interaction of sex with birth cohort and *ε* is the residual error subject to least-square minimization.

### Survival regression analysis

We evaluated the utility of *iLGS* in distinguishing differential longevity in the validation set. We used the Lifelines package^81^ (version 0.25.11) to fit a non-parametric Cox’s proportional hazard model according to the formula below:

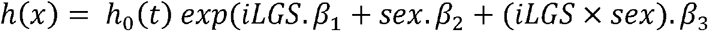

where, *h*_0_(*t*) is the baseline hazard estimated using Breslow’s method. To increase the stability of estimates we included a penalizer term of 0.0005 and L_1_-ratio of 1.0 to shrink the magnitude of 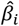 according to:

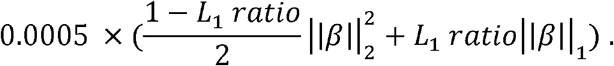

The significance of individual regression coefficients was investigated using the likelihood ratio test. The absence of deviation from proportional hazard assumptions was validated using the global test for Schoenfeld residuals. We also calculated the cumulative risk of death (as a function of age) across the five quintiles of iLGS based on the equation below:

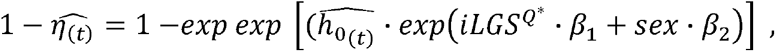

where 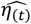 is the cumulative survival at the given time (*t*), 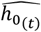 is the estimated baseline hazard at time t and *iLGS*^*Q*∗^ is the quintile of the iLGS score.

### Evaluation of the iLGS

We estimated survival curves across different quintiles of iLGS using the Kaplan-Meier estimator:

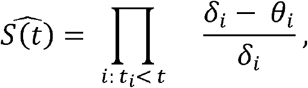

where, *θ*_*i*_ is the number of death events at a particular time *t*_*i*_, and *δ*_*i*_ is the fraction of the population at risk of death prior to time *t*_*i*_ . We investigated whether there is a significant difference in survival probability across quintiles of iLGS using the log-rank test.

We tested the performance of iLGS for distinguishing the “age at diagnosis” of three late-life cardiovascular pathologies (i.e., angina, arrhythmia, and interventional cardiac procedures) and three types of cancers (i.e., breast, prostate, and skin cancer) across 390 Ashkenazi Jewish individuals with available clinical and genotypic data. Kaplan-Meier survival curves across the top and bottom quintile of iLGS was compared using the log-rank test. We calculated the survival time post-diagnosis for each trait as the difference between the “age at death” and the “age at diagnosis”. In addition, we investigated the association of dummy *iLGS* quintiles with post-diagnosis medical intervention survival time using an ordinary least square regression while controlling for the gender effect.

### Validating iLGS performance in additional cohorts

We investigated the performance of iLGS for distinguishing survival in two additional cohorts. We used whole-genome sequences from 2,570 participants (male= 1,251; female= 1,319) of the Medical Genome Reference Bank (MGRB) cohort^70^. This cohort comprises healthy elderly individuals depleted for aging-related disorders and cancers. We derived the age of participants based on their respective dates of birth. At the time of analysis, the age of participants ranged from 75 to 102 years old (mean= 86; SD= 5.13). We corroborated participants’ reported sexes with X-chromosome heterozygosity estimates. Prior to the PRS calculation, we removed variant calls with read-depth (DP) ≤ 10 or FisherStrand bias (FS > 60). Furthermore, we excluded INDELs and multi-allelic variants and subjected the data to the standard QC procedure as explained above. This resulted in 8,496,911 autosomal SNPs to be used to compute iLGS. We also replicated the performance of iLGS across the 510 participants (194 males and 316 females) of the Wellderly cohort (**Supplementary Table 10**)^45^. Since these samples were sequenced using the Complete Genomics platform (Complete Genomics), we initially converted the “transvariants” calls to the conventional VCF format. We subsequently removed indels, multi-allelic SNPs, and variants with low read-depth DP ≤ 10 or variants flagged as “VQLOW”. Upon standard QC, 3,383,869 autosomal SNPs were used for iLGS calculation. Since age at death in these two cohorts was undetermined, we first binned participants of each cohort into three age categories: < 90, [90, 94], and ≥ 95. Then, we applied a two-sided Fisher’s exact test to investigate the association of top 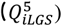 and bottom quintile *iLGS* 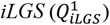 with age categories in each cohort independently.

### Proteomic analysis across quintiles

The proteomic assessment was carried out using the SomaScan assay V.4.0. Details related to the experimental design and quantification of the protein levels were described earlier^47^. Briefly, the relative concentration of 5,209 human-specific SOMAscan aptamers and 75 non-human and control aptamers were measured among 1,027 participants of the Einstein LonGenity cohort. Normalized aptamer concentrations in the relative fluorescent unit (RFU) were used to remove proteins and individuals with significant variation across array runs, according to Candia et al.^82^. Upon QC and removal of aptamers targeting non-human specific proteins, a total of 4,265 protein signals were obtained and used for proteomic analysis.

We quantified the association between the protein levels and the iLGS using a robust regression with MM-estimator for each gender group separately (123 males and 115 females). For protein targets tagged by multiple aptamers, only the most significant association signals were analyzed. To enhance the reliability of the regression in the presence of anomalous protein-levels, we used Cook’s distance to remove outliers with a residual value > 4/*n*. We carried out a gender-stratified comparison between the 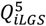 (top quintile iLGS) versus 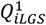 `(bottom quintile iLGS) across individuals in the ≥95 age bin, and applied the empirical Bayes moderated t-test using the Limma package`^83^ to compare normalized RFU intensities. All *P*-values were corrected for multiple hypothesis testing using the Benjamini-Hochberg method (FDR < 0.05). Significant proteins were used to search for potential drug candidates for repurposing opportunities in the DrugBank database (version 5.1.8)^55^. We restricted our analysis to drugs with clinical phase assignment as “experimental”, “investigational”, or “approved” status, corresponding to clinical trial phase I, II, and III, respectively.

## Data Availability

Summary statistics for the longevity association of analyzed variants in our Ashkenazi Jewish longevity cohort and SNP weights for polygenic scores derived in this study will be deposited on the lab website and will be available upon request. Due to privacy concerns for our research participants, individual-level genetic data from the Einstein longevity study are not publicly available; however, anonymized data will be shared upon request from a qualified academic investigator, providing the data transfer is approved by the Institutional Review Board and regulated by a material transfer agreement.
The whole-genome data from the Medical Genome Reference Bank and the Wellderly cohort is accessible through the European genome-phenome archive under study IDs EGAS00001003511 and EGAD00001003941, respectively. The genotyping array data for the GERA cohort (Resource for Genetic Epidemiology Research on Aging) is available through dbGaP Study Accession: phs000674.v2.p2.
We used genotypes and summary statistics from publicly available databases to estimate linkage disequilibrium correlations and construct polygenic scores. We used the genetic and clinical data from the UK Biobank under research project ID 58069 (https://www.ukbiobank.ac.uk/enable-your-research/approved-research/evolutionary-analysis-of-ageing-and-age-related-disorders-in-the-extant-human-population). The various summary statistics used to calculate genetic correlations are available from GeneAtlas (http://geneatlas.roslin.ed.ac.uk/), NealeLab (http://www.nealelab.is/uk-biobank) and GWASAtlas (https://atlas.ctglab.nl). We used DrugBank v.5.1.8 (https://go.drugbank.com/releases/5-1-8) to identify drugs with repurposing potential as gerotherapeutics.
Source data for figures in this study are available in the supplementary documents and upon request from the corresponding authors.

## Acknowledgments

This work was supported by funding to ZDZ from NIH Grants R01 AG057909, U19 AG056278, R01 AG061521, R01 AG057706, RF1 AG057341, R01 AG061155, P01 AG047200, P01 AG017242, P30 AG038072, and a Career Scientist Award from the Irma T. Hirschl Trust.

## Contributions

M.R.J. and Z.D.Z. conceived the formal analysis. M.R.J. executed the formal analysis. Z.D.Z., J.-R.L., J.M., Q.Z. and T.G. performed data curation. Z.W. performed variant imputation. M.K. performed summary statistics data munging. Z.D.Z. and N.B. conceived of the research goals and acquired funding. M.R.J. and Z.D.Z. wrote the original draft. J.V., N.B., S.M., G.A. and N.N. participated in the review and editing.

## Data availability

Summary statistics for the longevity association of analyzed variants in our Ashkenazi Jewish longevity cohort and SNP weights for polygenic scores derived in this study will be available upon request. Due to privacy concerns for our research participants, individual-level genetic data from the Einstein longevity study are not publicly available; however, anonymized data will be shared upon request from a qualified academic investigator, providing the data transfer is approved by the Institutional Review Board and regulated by a material transfer agreement. The whole-genome data from the Medical Genome Reference Bank and the Wellderly cohort is accessible through the European genome-phenome archive under study IDs EGAS00001003511 and EGAD00001003941, respectively. The genotyping array data for the GERA cohort (Resource for Genetic Epidemiology Research on Aging) is available through dbGaP Study Accession: phs000674.v2.p2. We used genotypes and summary statistics from publicly available databases to estimate linkage disequilibrium correlations and construct polygenic scores. We used the genetic and clinical data from the UK Biobank under research project ID 58069 (https://www.ukbiobank.ac.uk/enable-your-research/approved-research/evolutionary-analysis-of-ageing-and-age-related-disorders-in-the-extant-human-population). The various summary statistics used to calculate genetic correlations are available from GeneAtlas (http://geneatlas.roslin.ed.ac.uk/), NealeLab (http://www.nealelab.is/uk-biobank) and GWASAtlas (https://atlas.ctglab.nl). We used DrugBank v.5.1.8 to identify drugs with repurposing potential as gerotherapeutics which are available at (https://go.drugbank.com/releases/5-1-8). Source data for figures in this study are available in the supplementary documents and upon request from the corresponding author.

