## Extended figures for "Polygenic prediction of human longevity on the supposition of pervasive pleiotropy"

**Extended Figure 1:** A schematic overview of the analysis.

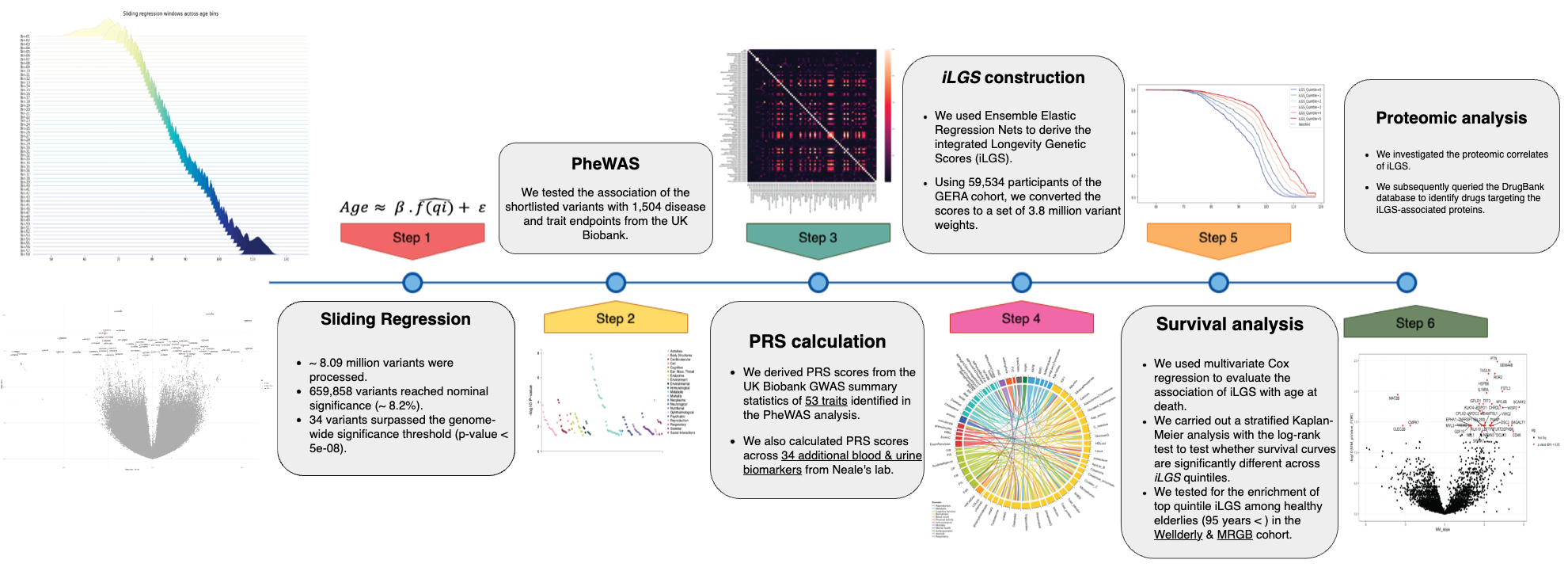

Our analysis constitutes six major steps; in **Step 01**, we used the sliding regression framework to track allele frequency changes across 48 age bins in the Einstein centenarian cohort. We fitted a robust linear regression with MM-estimator for each variant to derive the slope and p-value of association with chronological age. Variants surpassing the genome-wide significance threshold (p < 5e-8) were replicated in the UK Biobank; in **Step 02**, the association of 34 candidate variants with 614 disease endpoints and traits were investigated in a PheWAS analysis. For each variant, we used the Bonferroni-corrected association p-values for shortlisting associated traits; in **Step 03**, we used GWAS summary statistics across the 87 traits (primarily from the UK Biobank) to calculate the polygenic scores (PGS) for all participants in the Einstein centenarian cohort (n= 952); in **Step 04**, we split our cohort into the derivation set (n= 715) and validation set (n= 273). Using the derivation portion of our cohort, we applied a stacked Elastic-net regression framework to effectively combine PGS across the 87 traits and construct a composite prognostic score to distinguish survival chances. We trained the ensemble model on 65% of the derivation set and tested on the remaining 35%. Upon five-fold cross-validation, our model achieved an AUC of 0.87. Using coefficients derived from our stacked model, we computed the integrated Longevity Genetic Scores (iLGS). Subsequently, using an external cohort (GERA cohort), we converted the scores to a set of 3.8 million variant weights. In **Step 05**, we carried out survival analysis to test the performance of iLGS in predicting survival in the validation set as well as two external cohorts including the Wellderly and MRGB cohorts; in **Step 06**, we carried out a gender-stratified association analysis to identify proteomic correlates of iLGS. Proteins significantly associated with iLGS were subsequently queried in the DrugBank database to identify druggable targets. Drugs targeting or interacting with iLGS associated proteins were investigated for potential repurposing as senolytics.

**Extended Figure 2:** Overview of the iLGS model construction pipeline.

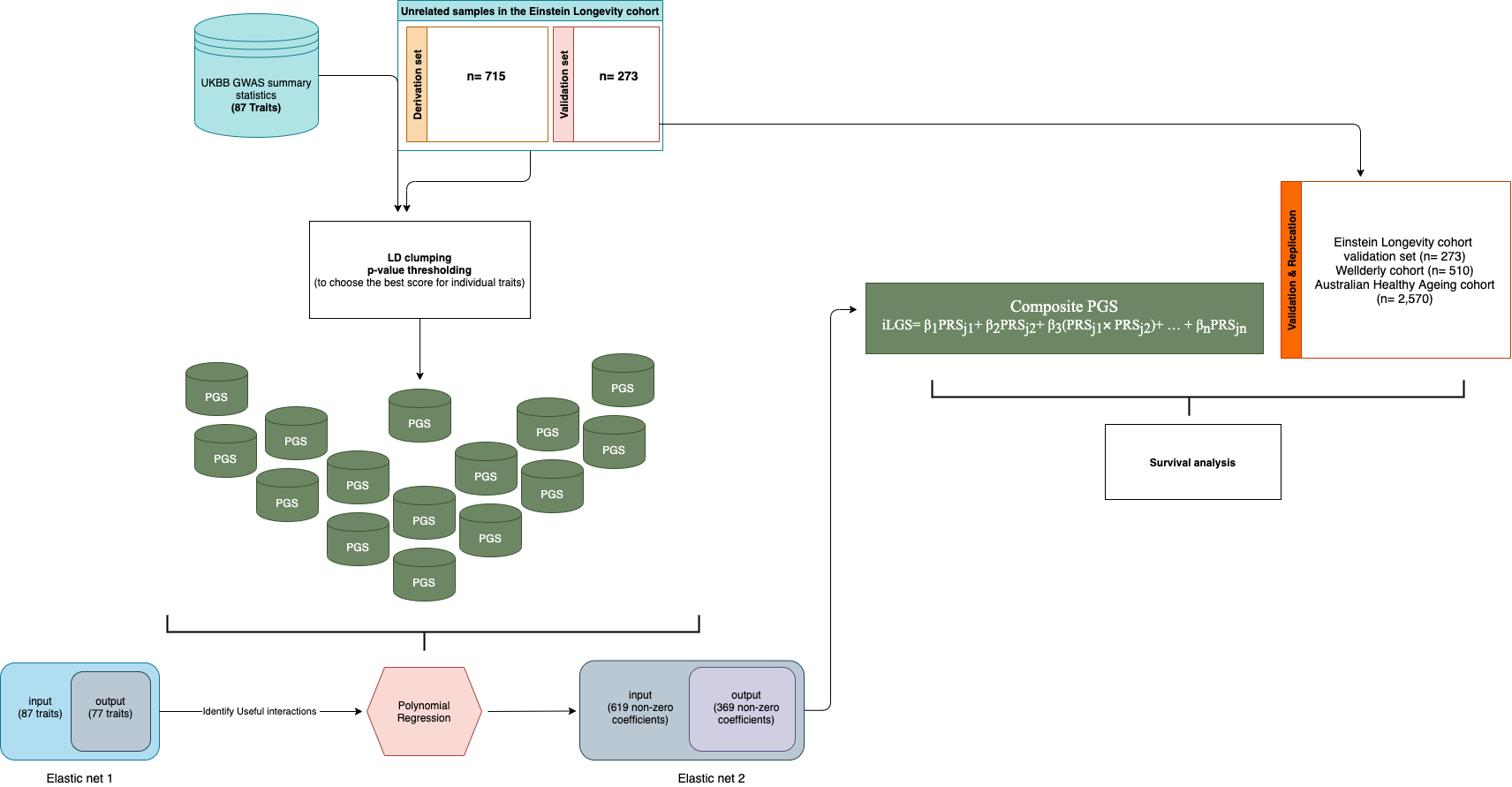

We derived integrated Longevity Genetic Scores (iLGS) in three steps; (a.) First, across unrelated individuals in the Einstein longevity cohort, we calculated polygenic scores for a total of 87 traits (53 associated traits from PheWAS analysis + 34 additional blood and urine biomarkers). We used LD clumping and p-value thresholding to derive the best score for each trait; (b.) Next, we applied a stacked Elastic-net regression framework to combine the polygenic scores and derive iLGS. This method essentially entails passing the output from one model to the next to increase model accuracy and derive meaningful insight. By sandwiching a polynomial regression between two Elastic-net regressions, we created a search space for the model to select the most informatic PGS and their interactions for predicting longevity. We started with 87 PGS as input and derived to 369 PGS and interaction terms with model accuracy around 91% upon five-fold cross-validation; (c.) Using the validation portion of the Einstein longevity cohort (n=273) and two additional cohorts, including the Wellderly (n= 510) & Australian Healthy Ageing cohort (MRGB; n=2,570) we replicated the predictive performance of iLGS for distinguishing differential lifespan.

**Extended Figure 3:** The interaction heatmap plot between the traits’ PGS across twelve major domains.

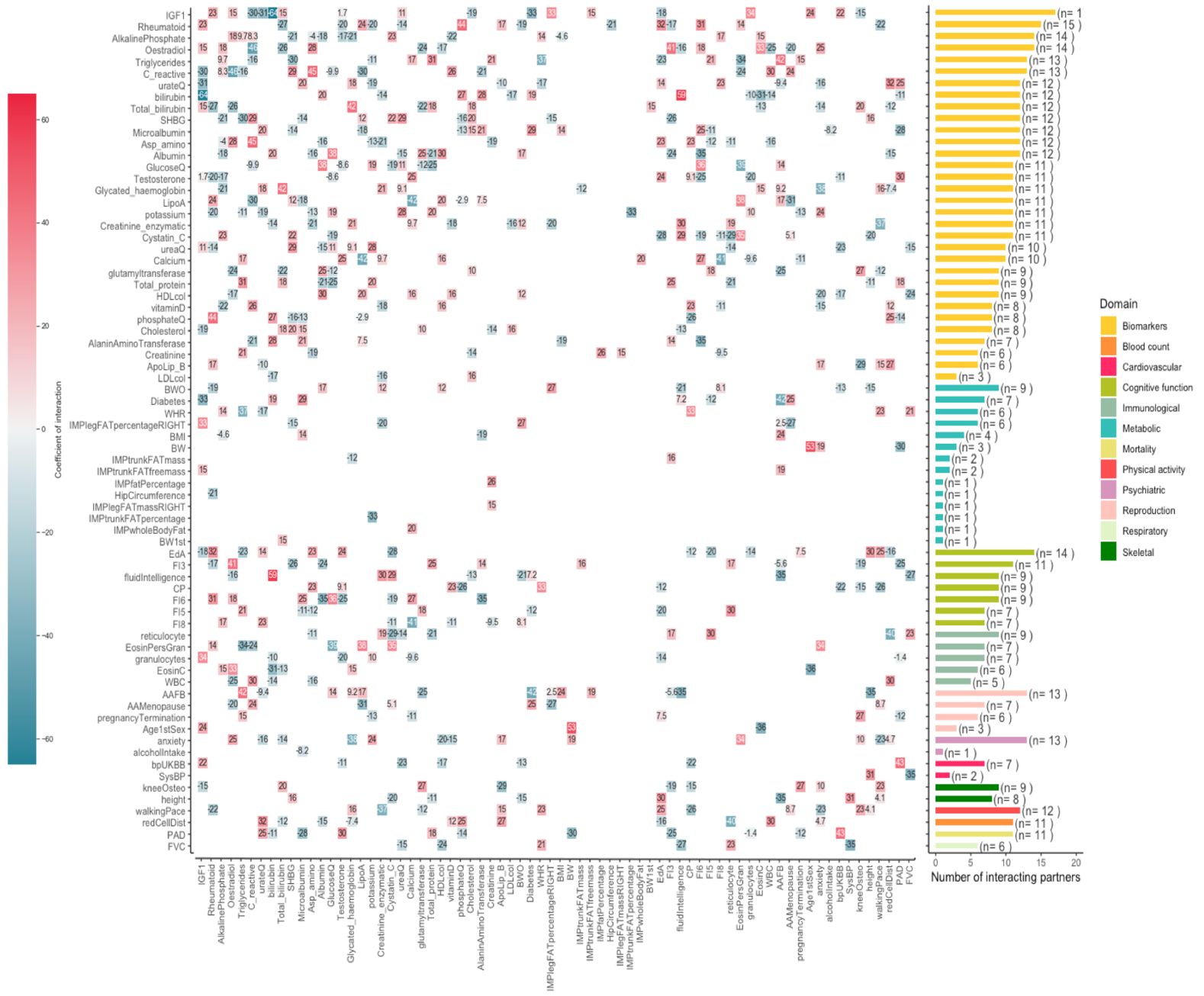

The heatmap plot is showing the interaction between the traits’ PGS used in constructing the iLGS. Interacting pairs with positive coefficients are shown in red, and pairs with negative coefficients in blue. The numbers inside the heatmap boxes correspond to the size of interacting pairs’ coefficients in the final model. The bar plot to the right shows the number of interacting pairs per trait PGS. The colour of each bar corresponds to the respective domain of the trait; (IGF1: Insulin growth factor-1, Rheumatoid: Rheumatoid factor, AlkalinePhosphate: Alkaline phosphatase, urateQ: Urate, bilirubin: Direct bilirubin, SHBG: Sex hormone binding globulin, Asp_amino: Aspartate aminotransferase, LipoA: Apolipoprotein A, Creatinine_enzymatic: Creatinine in urine (enzymatic), glutamyltransferase: Gamma glutamyltransferase, HDLcol: HDL cholesterol, AlaninAminoTransferase: Alanine aminotransferase, ApoLip_B: Apolipoprotein B, LDLcol: LDL cholesterol, BWO: Birth weight of offspring, BW: Birth weight, BW1st: Birth weight of first offspring, EdA: Educational attainment; FI3: Fluid intelligence-word interpolation; CP: Cognitive performance, FI6: Fluid intelligence- conditional arithmetic, FI5: Fluid intelligence- family relationship calculation, FI8: Fluid intelligence- chained arithmetic, reticulocyte: Immature reticulocyte fraction, EosinPersGran: Eosinophil percentage of granulocytes, granulocytes: Neutrophil percentage of granulocytes, EosinC: Eosinophil count, WBC: White blood cell count, AAFB: Age at first live birth, AAMenopause: Age at menopause, Age1stSex: Age first had sexual intercourse, bpUKBB: Hypertension, SysBP: Systolic blood pressure, kneeOsteo: Knee Osteoarthritis, redCellDist: Red cell (erythrocyte volume) distribution width, PAD: Parents’ age at death, FVC: Forced vital capacity (FVC).

**Extended Figure 4:** Cumulative incidence of death by iLGS quintile.

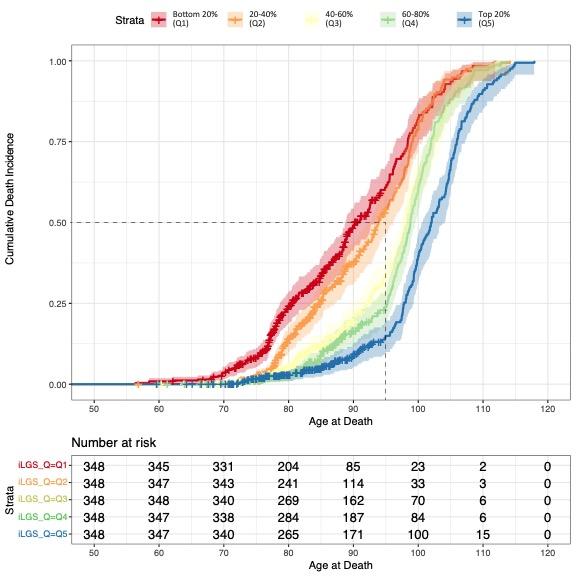

Shown is the cumulative incidence of death across different quintiles of iLGS. Across the bottom two quintiles ($Q_{iLGS}^{1}, Q_{iLGS}^{2}$), the cumulative incidence of death rapidly picks up after the age of 70, and more than 50% of the population fail to reach the age of 95. In comparison across the top two quintiles ($Q_{iLGS}^{1}, Q_{iLGS}^{2}$), the rate of death significantly picks up after the age of 95. By the age of 100, over 75% of the individuals in the bottom quintiles ($Q_{iLGS}^{1}, Q_{iLGS}^{2}$) are dead, while more than 50% of the individuals in the top quintile ($Q_{iLGS}^{5}$) are still alive.
