## Supplementary figures for "Polygenic prediction of human longevity on the supposition of pervasive pleiotropy"

**Supplementary Figure 1:** The age distribution and respective normal quantile-quantile plot for the Einstein longevity cohort.

a.

b.

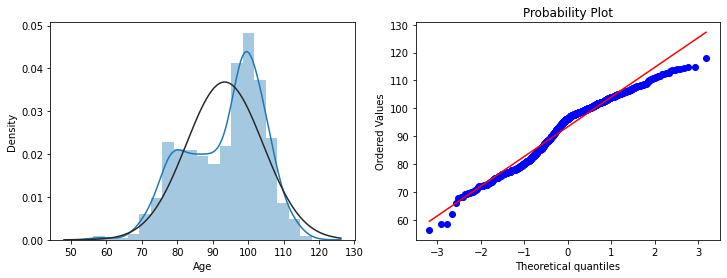

**a.** Histogram of age distribution across the 957 participants of Einstein longevity cohort. The black curve identifies the theoretical normal distribution. **b**. Normal quantile-quantile plot showing the right tail of the age-distribution in Einstein cohort is slightly lighter than the theoretical normal distribution (Q _empirical_ < Q _theoretical_).

**Supplementary Figure 2:** Volcano plot representing the associated variants from the sliding regression framework.

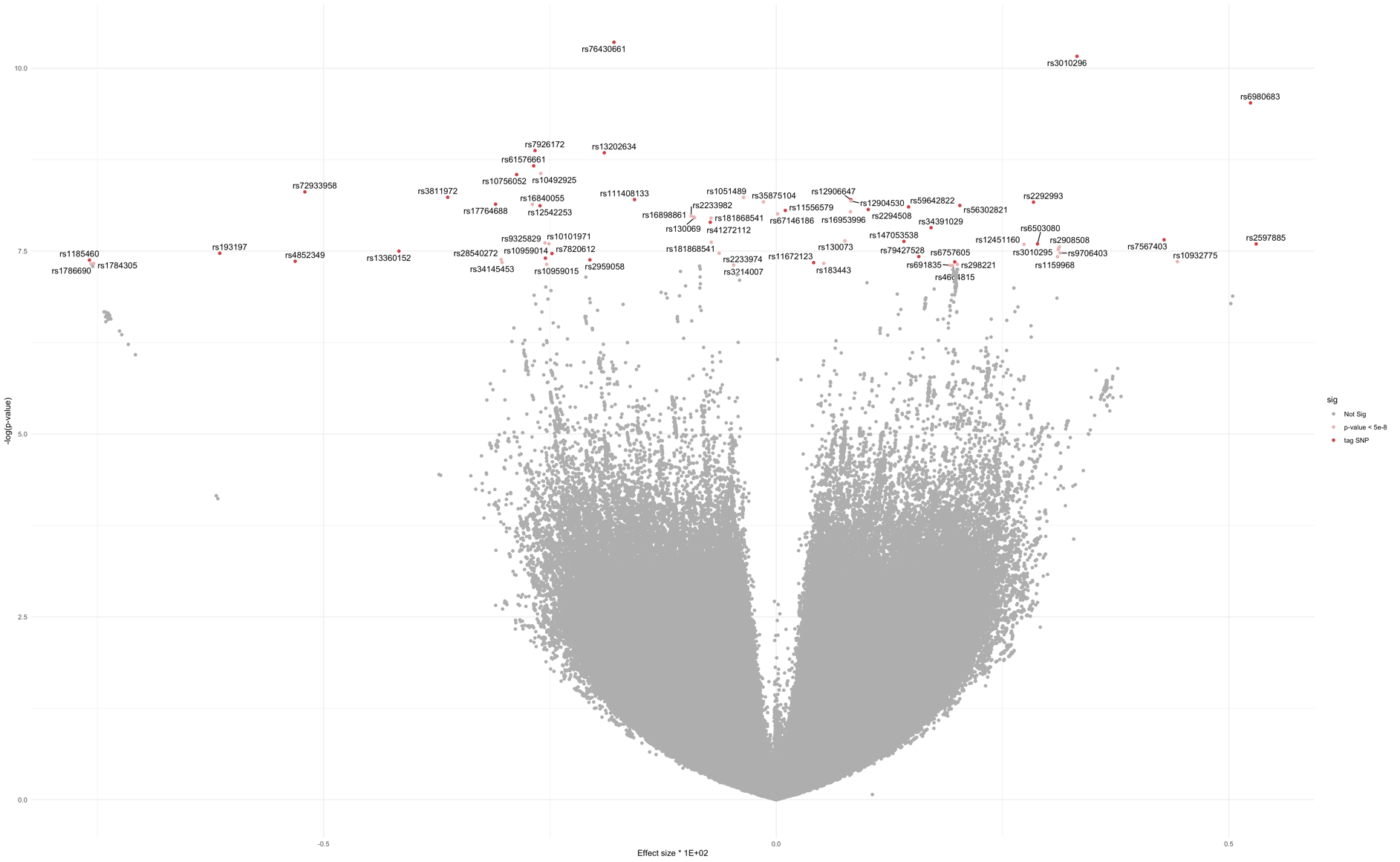

The x-axis in this volcano representation of results corresponds to the slope of regression. The pro-longevity variants whose frequency increases by age are clustered to the right of the plot and pro-ageing variants with negative slopes to the left. The y-axis shows the p-values in logarithmic scales with genome-wide significant variants identified in red. Variants’ coordinates are defined according to the GRCh37/hg19 assembly.

**Supplementary Figure 3:** *APOE* haplotypes frequency trajectories with age bins in the Einstein centenarian cohort.

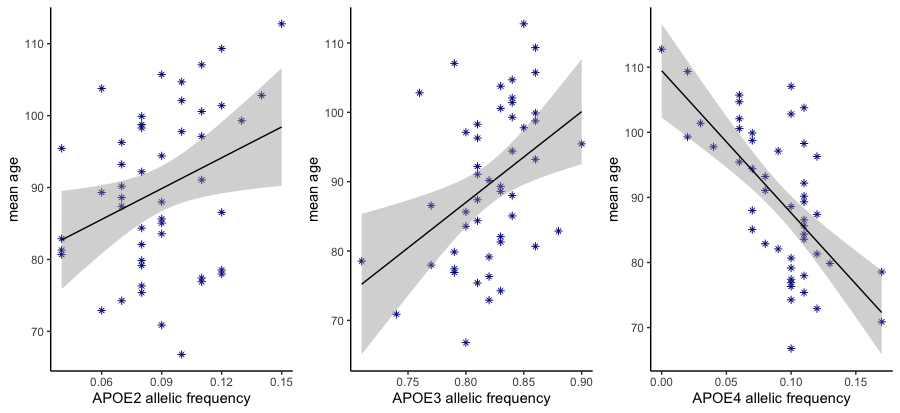

*R^2^* = 0.41

β= -21.84, *p* = 4.25e-07

*R^2^* = 0.08

β= 14.28, *p* = 2.44e-02

*R^2^* = 0.15

β= 13.09, *p* = 4.17e-03

The frequency of *APOE* haplotypes tagged by rs429358 and rs7412 were calculated in each age bin and regressed onto the mean age of each bin. Consistent with the previously reported deleterious effect of *APOE*-ε_4_ haplotype on longevity, we identified a strong negative effect of *APOE*-ε_4_ haplotype on lifespan. We also identified a nominally significant positive association *APOE*-ε_2_ & ε_3_ haplotypes with lifespan, but the association p-value for *APOE*-ε_2_ does not withstand the multiple testing correction. Further, the higher frequencies of *APOE*-ε_3_ haplotype among elderly age-bins can be explained by the relative depletion of *APOE*-ε_4_ haplotype in these age groups and cannot be independently accounted for the increased longevity in these age groups.

**Supplementary Figure 4:** Chromatin interactions and eQTLs of the lead SNPs on chromosome 5.

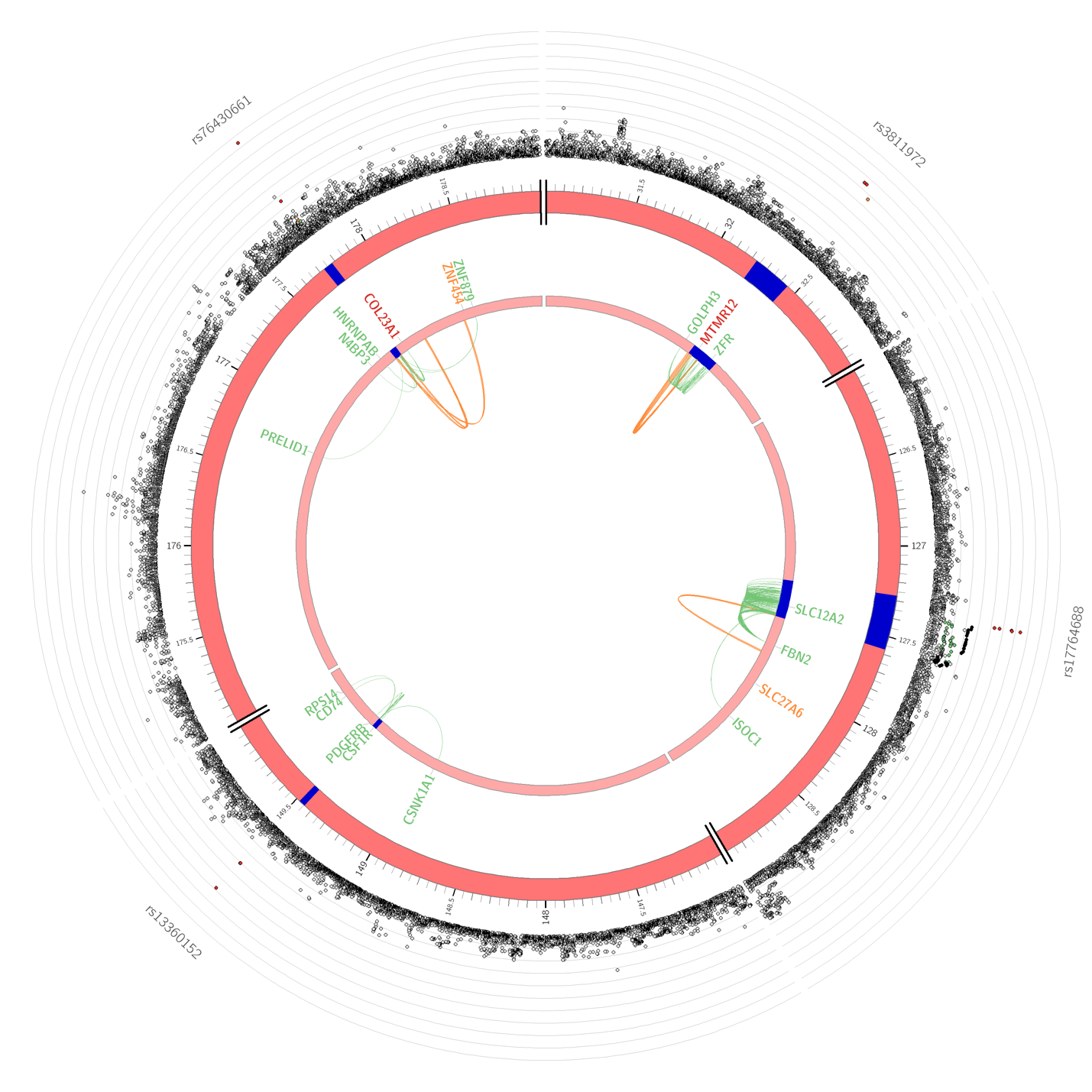

Among the 34 variants identified through the sliding regression framework, rs76430661 on 5q35.3 shows the strongest association signal. The outer layer is the Manhattan plot showing the association *P*-value of variants on chromosome 5. Candidate variants within each risk locus are colored according to their respective $r^{2}$ with the lead SNP (red: $r^{2}> 0.8$, orange: $r^{2}> 0.6$ and green:$r^{2}> 0.2$) according to the 1000 genomes phase 3 reference panel (EUR population). Risk loci surrounding the independent SNPs are highlighted in blue. Green and orange chord diagrams in the inner circle identify genes prioritized by eQTL and chromatin interactions, respectively. Genes prioritized by both eQTL and chromatin interaction mapping are highlighted in red.

**Supplementary Figure 5:** Chromatin interactions and eQTLs of the lead SNPs on chromosome 6.

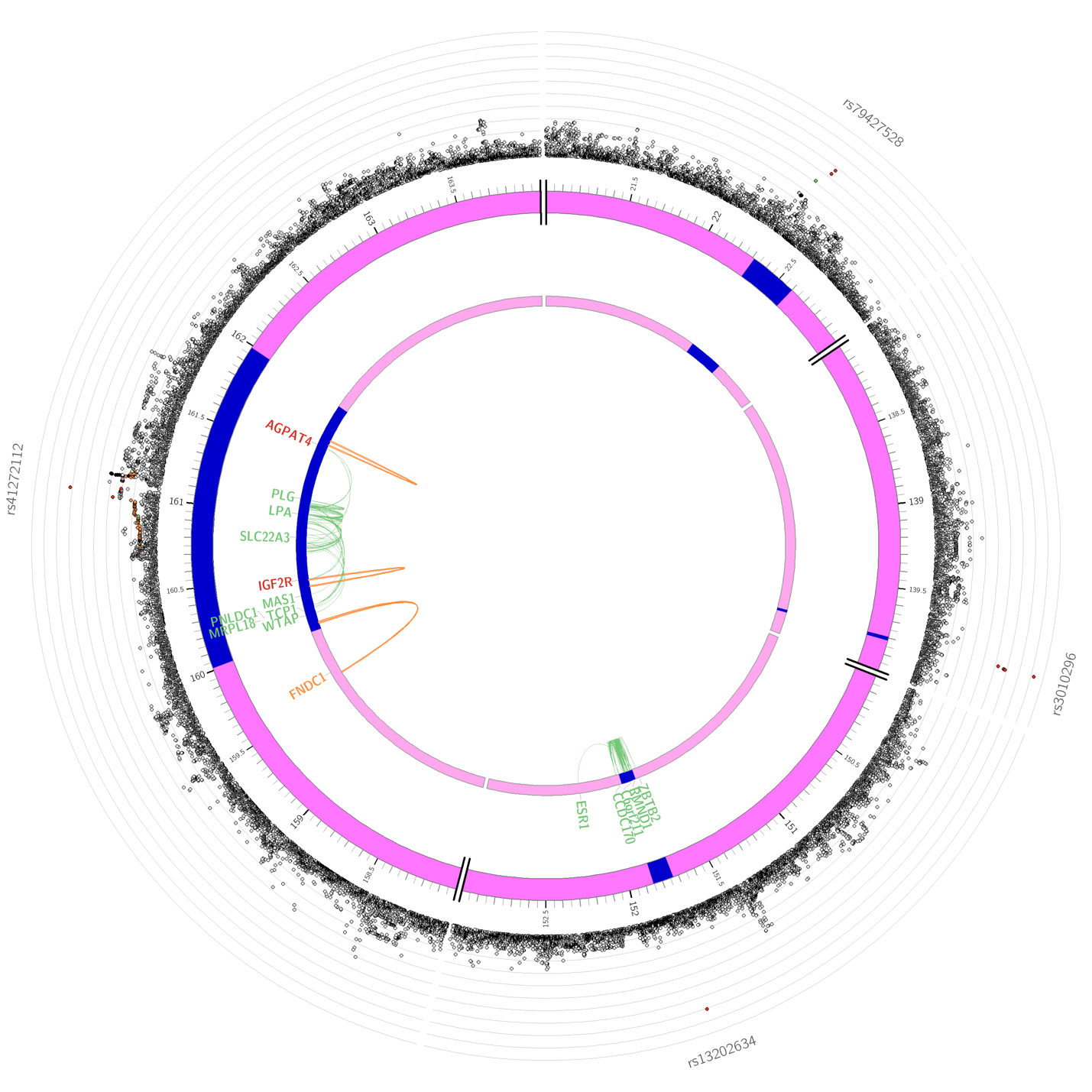

The risk locus surrounding rs41272112 on 6q26 highlights one of the two risk loci with the highest density of prioritized genes that are physically located in the risk locus. The outer layer is the Manhattan plot showing the association *P*-value of variants on chromosome 5. Candidate variants within each risk locus are colored according to their respective $r^{2}$ with the lead SNP (red: $r^{2}> 0.8$, orange: $r^{2}> 0.6$ and green:$r^{2}> 0.2$) according to the 1000 genomes phase 3 reference panel (EUR population). Risk loci surrounding the independent SNPs are highlighted in blue. Green and orange chord diagrams in the inner circle identify genes prioritized by eQTL and chromatin interactions, respectively. Genes prioritized by both eQTL and chromatin interaction mapping are highlighted in red.

**Supplementary Figure 6:** Chromatin interactions and eQTLs of the lead SNPs on chromosome 19.

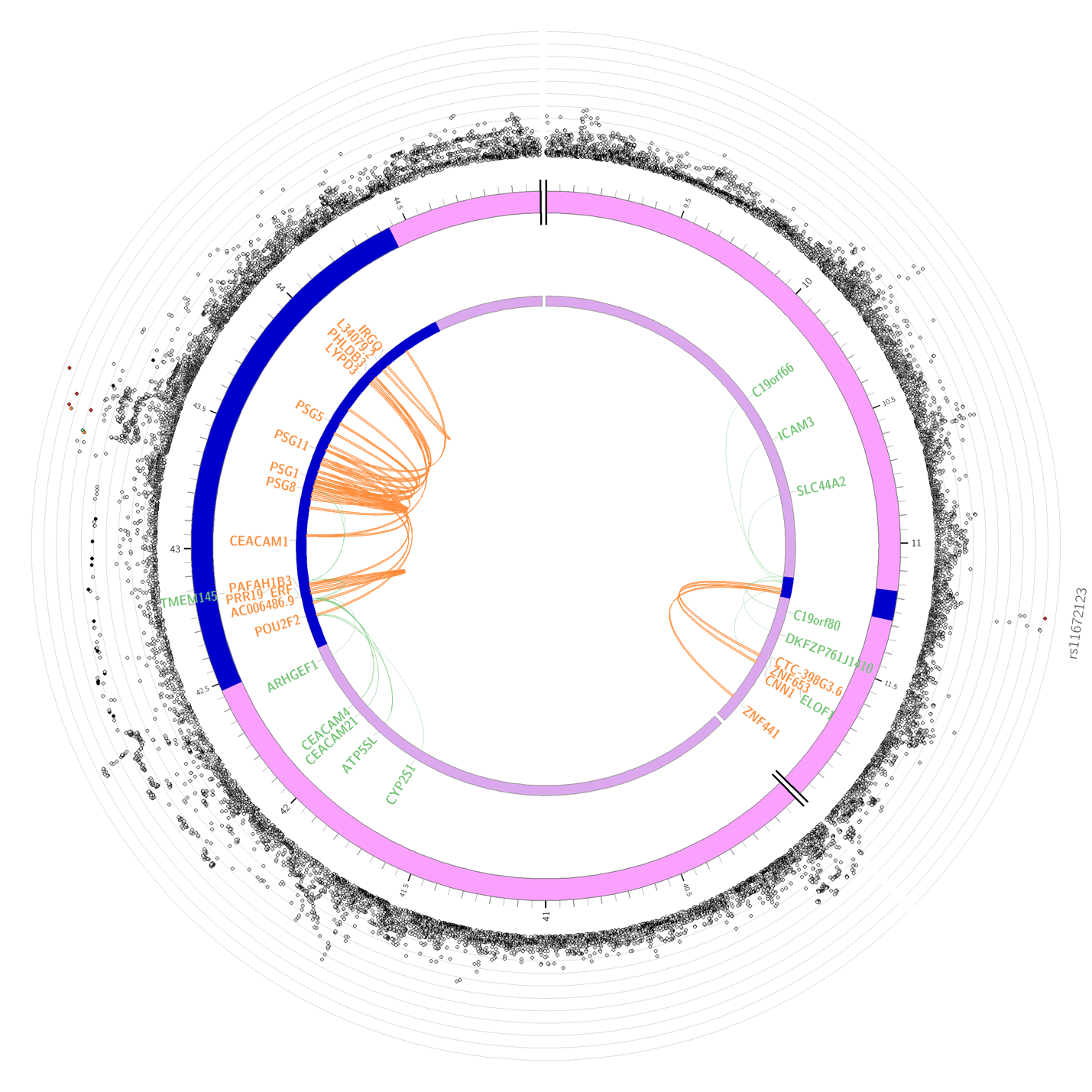

The risk locus on 19q13 surrounding rs147053538 highlights the second risk locus with the highest density of prioritized genes physically located in the region. The outer layer is the Manhattan plot showing the association *P*-value of variants on chromosome 5. Candidate variants within each risk locus are colored according to their respective $r^{2}$ with the lead SNP (red: $r^{2}> 0.8$, orange: $r^{2}> 0.6$ and green:$r^{2}> 0.2$) according to the 1000 genomes phase 3 reference panel (EUR population). Risk loci surrounding the independent SNPs are highlighted in blue. Green and orange chord diagrams in the inner circle identify genes prioritized by eQTL and chromatin interactions, respectively. Genes prioritized by both eQTL and chromatin interaction mapping are highlighted in red.

**Supplementary Figure 7:** Chromatin interactions and eQTLs of the lead SNP on chromosome 9.

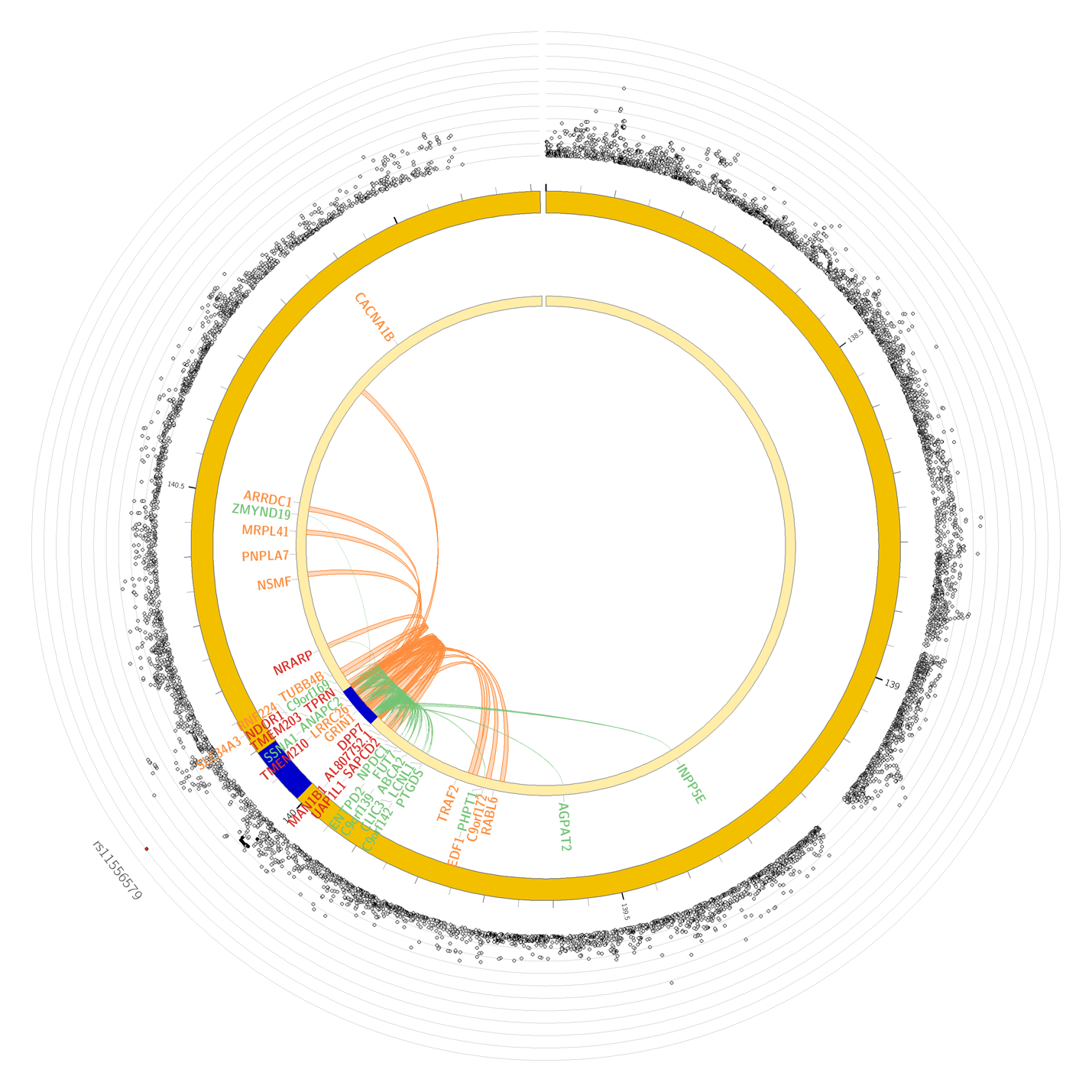

The risk locus surrounding rs11556579 on 9q34, highlights the most prolific regulatory activity with the highest number of genes prioritised by both eQTL and chromatin interaction mapping (n= 43). The outer layer is the Manhattan plot showing the association *P*-value of variants on chromosome 5. Candidate variants within each risk locus are colored according to their respective $r^{2}$ with the lead SNP (red: $r^{2}> 0.8$, orange: $r^{2}> 0.6$ and green:$r^{2}> 0.2$) according to the 1000 genomes phase 3 reference panel (EUR population). Risk loci surrounding the independent SNPs are highlighted in blue. Green and orange chord diagrams in the inner circle identify genes prioritized by eQTL and chromatin interactions, respectively. Genes prioritized by both eQTL and chromatin interaction mapping are highlighted in red.

**Supplementary Figure 8:** Chromatin interactions and eQTLs of the lead SNP on chromosome 2.

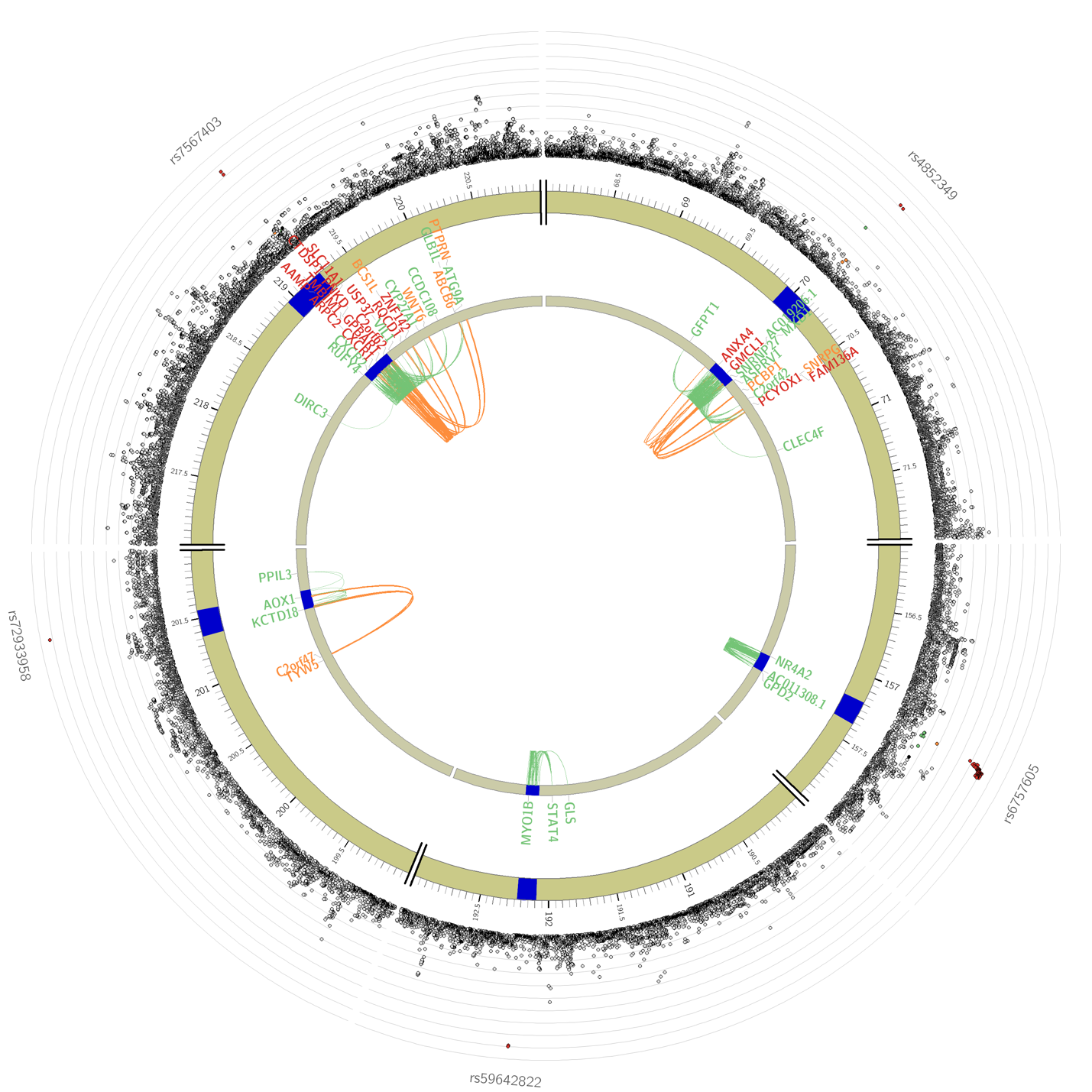

The two risk loci on 2q24 and 2q32 (surrounding rs6757605 and rs59642822) highlighting the eQTLs for *GPD2* and *GLS*, respectively. Downregulation of both genes have shown to increase lifespan in animal models. The outer layer is the Manhattan plot showing the association *P*-value of variants on chromosome 5. Candidate variants within each risk locus are colored according to their respective $r^{2}$ with the lead SNP (red: $r^{2}> 0.8$, orange: $r^{2}> 0.6$ and green:$r^{2}> 0.2$) according to the 1000 genomes phase 3 reference panel (EUR population). Risk loci surrounding the independent SNPs are highlighted in blue. Green and orange chord diagrams in the inner circle identify genes prioritized by eQTL and chromatin interactions, respectively. Genes prioritized by both eQTL and chromatin interaction mapping are highlighted in red.

**Supplementary Figure 9:** Schematic representation of interactions between traits’ PGS in the iLGS model.

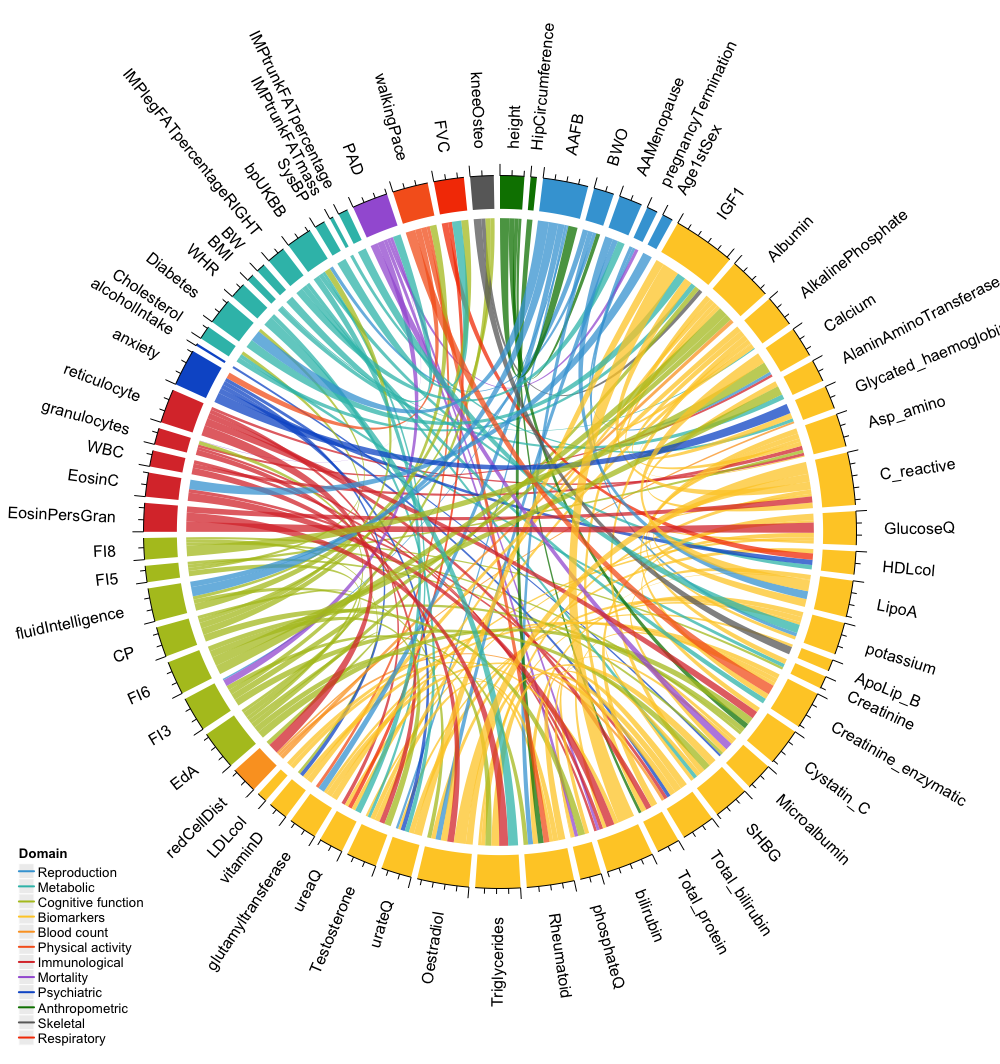

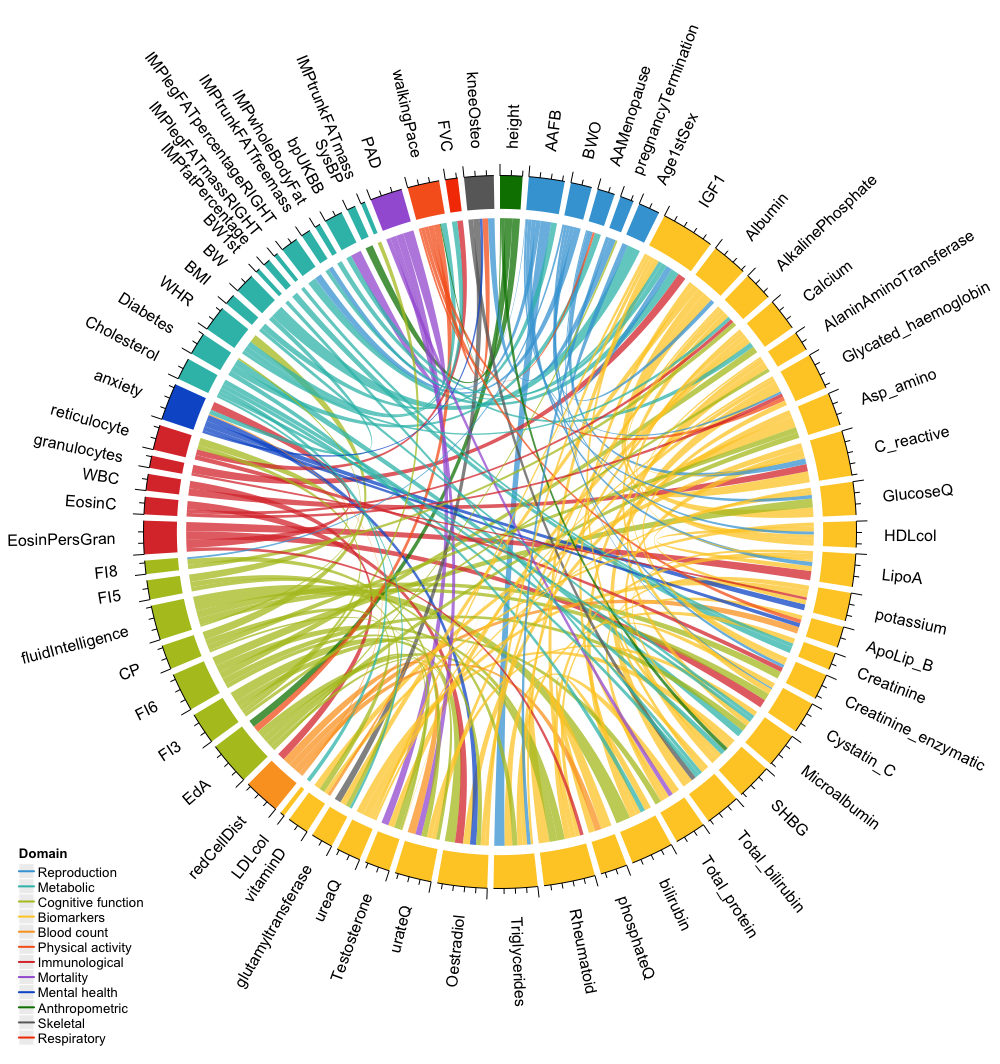

a.

b.

Chord diagrams showing the pattern of interaction between PGS of traits with positive (**S4. A**) and negative (**S4. B**) coefficients in the iLGS model across the 12 major domains. The chords’ thickness is proportional to the absolute size of the interaction coefficient in the final model. (IGF1: Insulin growth factor-1, Rheumatoid: Rheumatoid factor, AlkalinePhosphate: Alkaline phosphatase, urateQ: Urate, bilirubin: Direct bilirubin, SHBG: Sex hormone binding globulin, Asp_amino: Aspartate aminotransferase, LipoA: Apolipoprotein A, Creatinine_enzymatic: Creatinine in urine (enzymatic), glutamyltransferase: Gamma glutamyltransferase, HDLcol: HDL cholesterol, AlaninAminoTransferase: Alanine aminotransferase, ApoLip_B: Apolipoprotein B, LDLcol: LDL cholesterol, BWO: Birth weight of offspring, BW: Birth weight, BW1st: Birth weight of first offspring, EdA: Educational attainment; FI3: Fluid intelligence-word interpolation; CP: Cognitive performance, FI6: Fluid intelligence- conditional arithmetic, FI5: Fluid intelligence- family relationship calculation, FI8: Fluid intelligence- chained arithmetic, reticulocyte: Immature reticulocyte fraction, EosinPersGran: Eosinophil percentage of granulocytes, granulocytes: Neutrophil percentage of granulocytes, EosinC: Eosinophil count, WBC: White blood cell count, AAFB: Age at first live birth, AAMenopause: Age at menopause, Age1stSex: Age first had sexual intercourse, bpUKBB: Hypertension, SysBP: Systolic blood pressure, kneeOsteo: Knee Osteoarthritis, redCellDist: Red cell (erythrocyte volume) distribution width, PAD: Parents’ age at death, FVC: Forced vital capacity (FVC).

**Supplementary Figure 10:** Manhattan plot showing the genome-wide association of SNPs with iLGS.

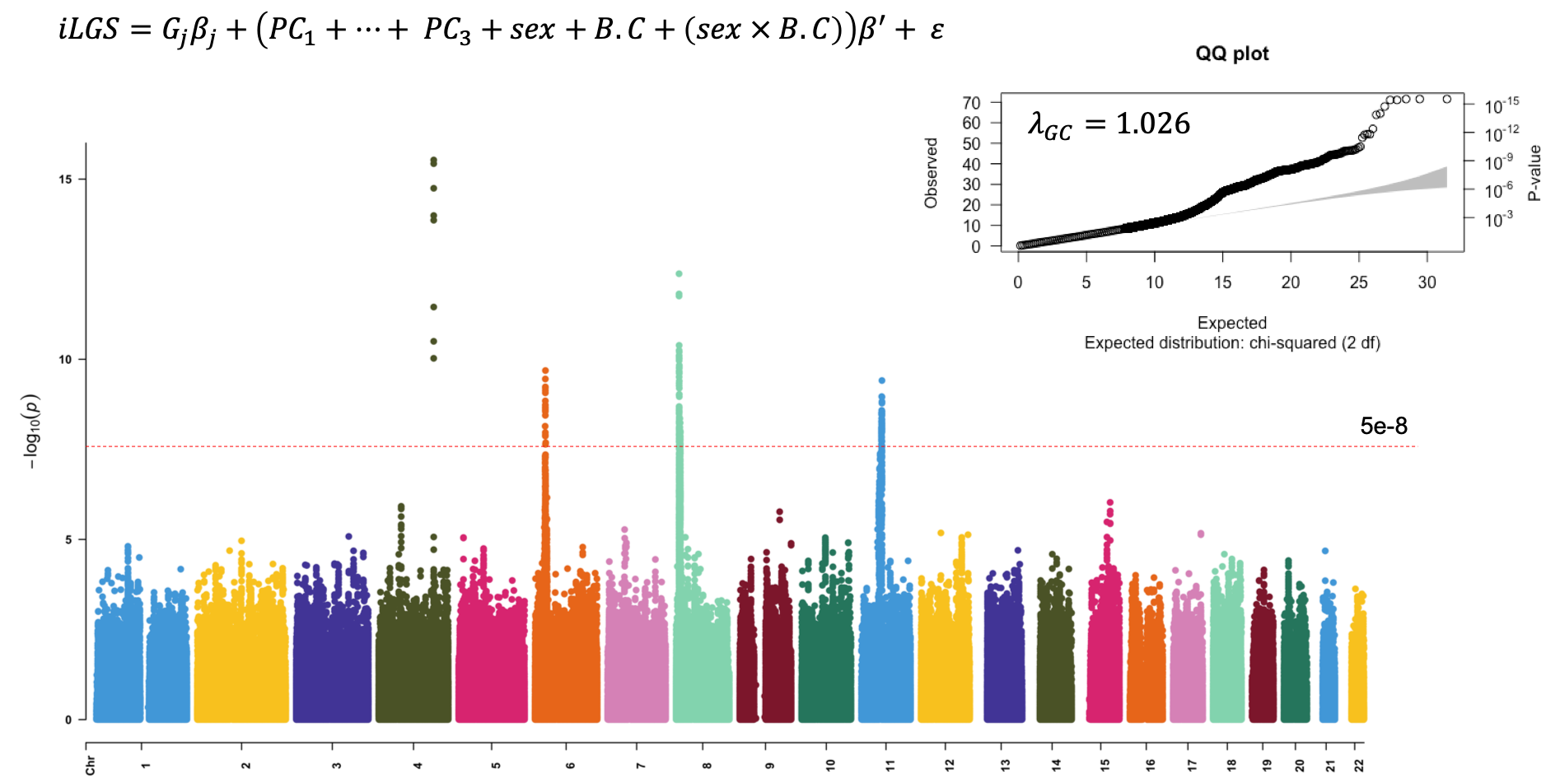

We used a subsample of the GERA cohort with European ancestry (n= 62, 268) to derive the SNP weights. We used a generalized linear formwork to derive iLGS SNP weights according to the formula specified above. The quantile-quantile (Q-Q) plot of association and the magnitude of genomic-control ($\lambda_{GC}=1.026$) confirm the absence of confounding in the derivation of SNP weights. The Q-Q plot shows the deviation of the observed association p-values against expected p-values under the null model of no significance. Since the deviation is observed exclusively in the right tail of p-value distribution, we concluded that the inflation of test statistics is primarily due to the polygenic inheritance of iLGS (i.e. longevity)^1^.

**Supplementary Figure 11:** Plot depicting the association of top quintile iLGS ($Q_{iLGS}^{5}$) with extreme obesity (BMI> 40 kgm^-2^) and treatment history for high blood pressure and high cholesterol in the MRGB cohort.

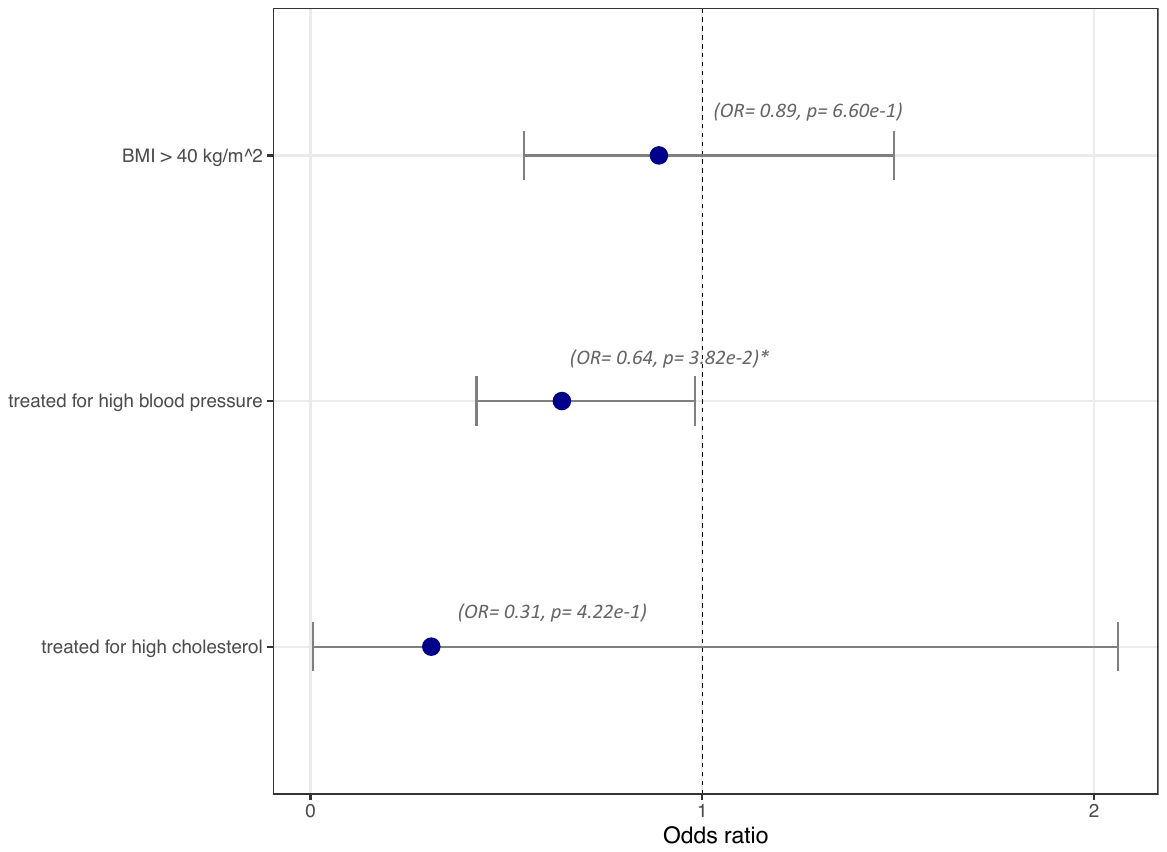

Using the available data in the MRGB cohort, we investigated the association of top quintile iLGS with three age-related traits. Among individuals with extreme obesity or treatment history for high blood pressure or high cholesterol, top quintile iLGS is uncommon (OR < 1). However, this depletion is only statistically significant for treatment history for high blood pressure (asterisks identify significant associations).

**Supplementary Figure 12:** Box and Whisker plot illustrating the burden of pathogenic rare variants across the top and bottom quintile iLGS in the Centenarian cohort.

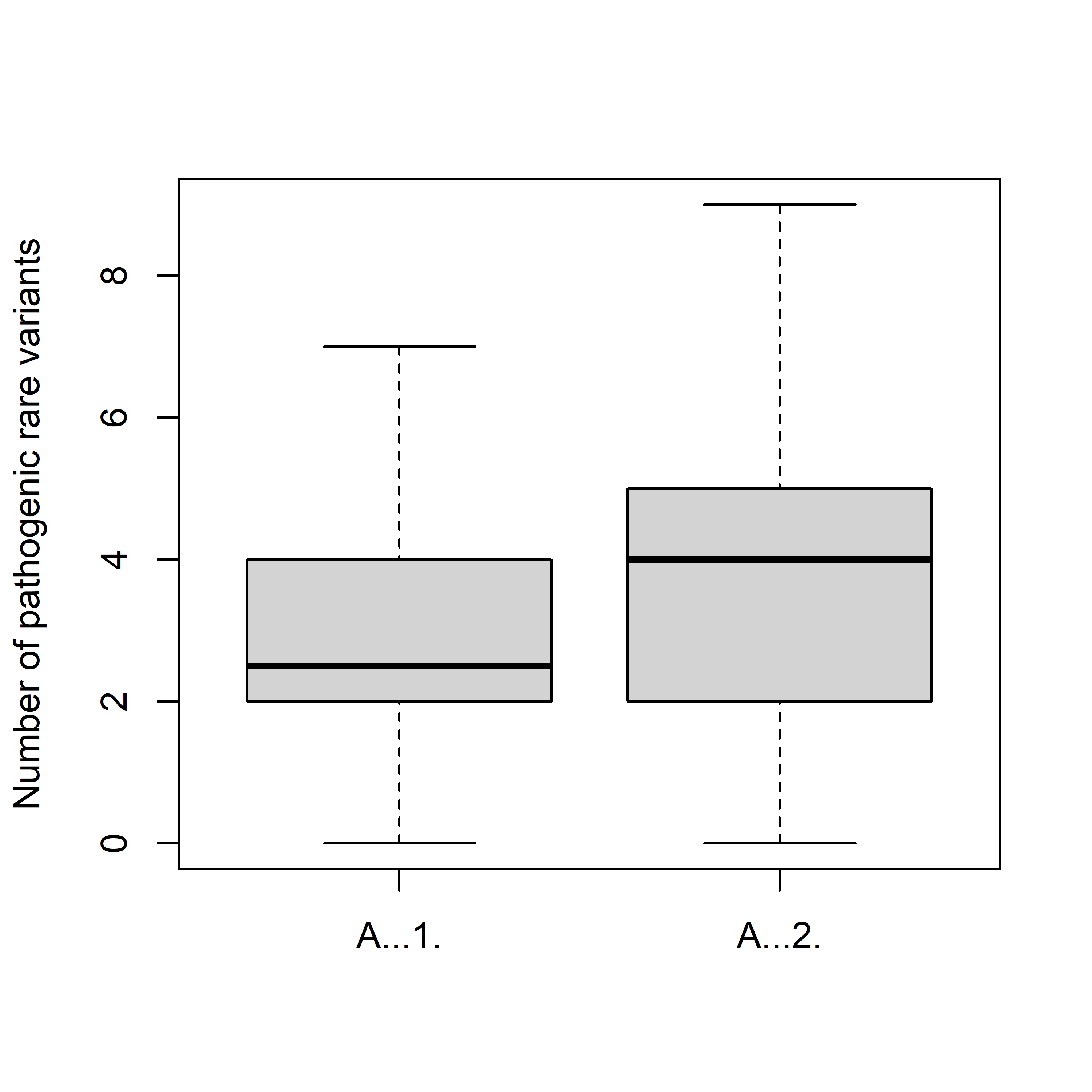

93 top Q centenarian

52 bottom Q centenarian

*P* = 0.027

**Supplementary Figure 13:** Estimates of lifespan narrow-sense heritability ($h^{2}$) partitioned by chromosome.

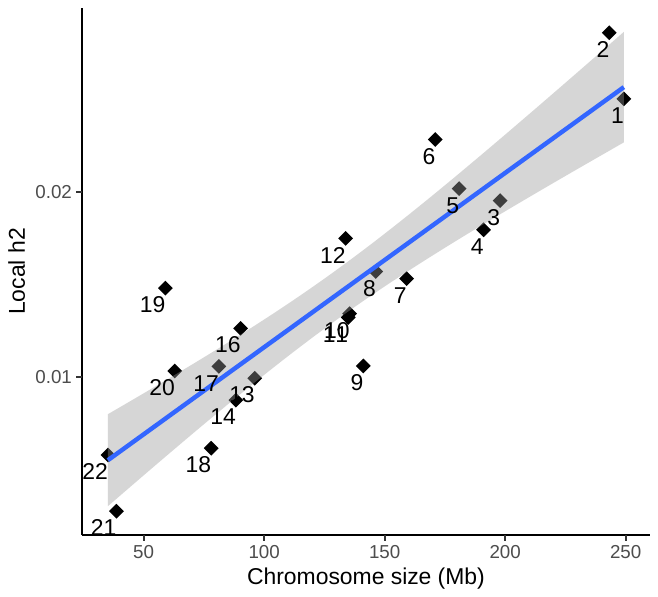

The estimates of narrow-sense heritability ($h^{2}$) is correlated with the length of autosomes. This indicates that genetic determinants of lifespan are thinly distributed along the length of the genome.

**Supplementary Figure 14:** The age distribution and respective normal quantile-quantile plot for the MRGB cohort.

a. ASPREE sub-cohort

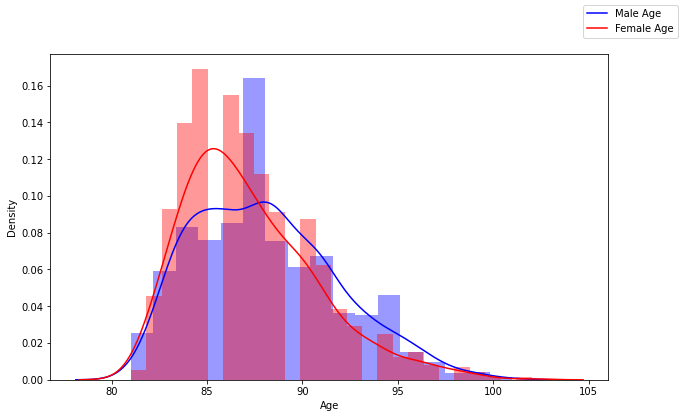

b. 45 and Up sub-cohort

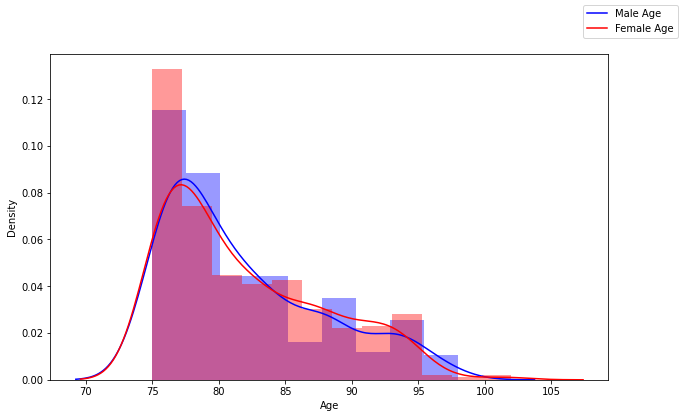

The MRGB cohort is comprised of two sub-cohorts. The age distribution for participants who are consented through the ASPirin in Reducing Events in the Elderly (ASPREE) clinical trial (Monash University, Melbourne) is shown in panel a and for those consented through the 45 and Up study (Sax Institute, Sydney) is shown in panel b. All participants are free from cardiovascular disease, degenerative neurological disorders and, cancer.
