## Supplemental Table 10 for "Polygenic prediction of human longevity on the supposition of pervasive pleiotropy"

**Supplementary Table 10.** Demographic characteristics of cohorts used in the study

|  | **Einstein longevity cohort (n= 957)** | | | |
| --- | --- | --- | --- | --- |
|  | Male | | Female | |
| Number of subjects (%) | 356 (37.19%) | | 601 (62.80%) | |
| Age at death, years  (mean ± SD) | 90.94 (±10.47) | | 94.81 (±10.80) | |
|  | **^+^Wellderly cohort (n= 510)** | | | |
|  | Male | | Female | |
| Number of subjects | 194 (38.03%) | | 316 (61.96%) | |
| Age at recruitment, years  (mean ± SD) | 86.06 (±4.67) | | 86.45(±5.04) | |
|  | **MRGB cohort (n= 2,570)** | | | |
|  | Male | | Female | |
| Number of subjects | 1,251 (48.68%) | | 1,319 (51.32%) | |
| Age at recruitment, years  (mean ± SD) | 86.64 (± 5.12) | | 85.54 (± 5.09) | |
|  | **UK Biobank (n= 20,890)** | | | |
|  | Male | | Female | |
| Number of subjects | 12,597 (60.30%) | | 8,293 (39.70%) | |
| Age at last contact, years  (mean ± SD) | 70.96 (± 5.86) | | 70.80 (±6.05) | |
|  | **^++^GERA cohort (n= 62,268)** | | | |
|  | Male | | Female | |
| Number of subjects | 24,950 (40.06%) | | 37,318 (59.93%) | |
| Birth year category | <= 1923 | 2,736 | <= 1923 | 3,172 |
|  | 1924- 1928 | 2,868 | 1924- 1928 | 3,252 |
|  | 1929- 1933 | 3,631 | 1929- 1933 | 4,467 |
|  | 1934- 1938 | 4,292 | 1934- 1938 | 5,696 |
|  | 1939- 1943 | 4,148 | 1939- 1943 | 6,233 |
|  | 1944- 1948 | 2,862 | 1944- 1948 | 4,831 |
|  | 1949- 1953 | 1,875 | 1949- 1953 | 3,536 |
|  | 1954- 1958 | 1,137 | 1954- 1958 | 2,371 |
|  | 1959- 1963 | 616 | 1959- 1963 | 1,432 |
|  | 1964- 1968 | 385 | 1964- 1968 | 1,085 |
|  | 1969- 1973 | 212 | 1969- 1973 | 639 |
|  | 1974- 1978 | 94 | 1974- 1978 | 327 |
|  | 1979- 1983 | 94 | 1979- 1983 | 277 |

+ The age of individuals over 90 in the Wellderly cohort is reported in bins with four year increments. Therefore, to calculate the mean age over the whole cohort, we used the mean of the age bins for all individuals in that age bin.

++ In the GERA cohort, the year of birth is censored, and only birth year categories are reported.
